# Influence of meteorological factors and drought on coccidioidomycosis incidence in California, 2000–2020

**DOI:** 10.1101/2022.02.03.22270412

**Authors:** Jennifer R. Head, Gail Sondermeyer-Cooksey, Alexandra K. Heaney, Alexander T. Yu, Isabel Jones, Abinash Bhattachan, Simon Campo, Robert Wagner, Whitney Mgbara, Sophie Phillips, Nicole Keeney, John Taylor, Ellen Eisen, Dennis P. Lettenmaier, Alan Hubbard, Gregory S. Okin, Duc J. Vugia, Seema Jain, Justin V. Remais

## Abstract

**Background:** Coccidioidomycosis is an emerging infection in the southwestern United States. We examined the effects of precipitation and temperature on the incidence of coccidioidomycosis in California during 2000-2020, and estimated incident cases attributable to the California droughts of 2007-09 and 2012-15.

**Methods:** We analyzed monthly California coccidioidomycosis surveillance data from 2000–2020 at the census tract-level using generalized additive models. Models included distributed lags of precipitation and temperature within each endemic county, pooled using fixed-effects meta-analysis. An ensemble prediction algorithm of incident cases per census tract was developed to estimate the impact of drought on expected cases.

**Results:** Across 14 counties examined, coccidioidomycosis was strongly suppressed during, and amplified following, the 2007-2009 and 2012-2015 droughts. An estimated excess of 1,358 and 2,461 drought-attributable cases were observed in California in the two years following the 2007-2009 and 2012-2015 droughts, respectively. These post-drought excess cases more than offset the drought-attributable declines of 1,126 and 2,192 cases, respectively, that occurred during the 2007-2009 and 2012-2015 droughts. Across counties, a temperature increase from the 25^th^ to 75^th^ percentile (interquartile range) in the summer was associated with a doubling of incidence in the following fall (incidence rate ratio (IRR): 2.02, 95% CI: 1.84, 2.22), and a one IQR increase in precipitation in the winter was associated with 1.45 (95% CI: 1.36, 1.55) times higher incidence in the fall. The effect of winter precipitation was stronger (interaction coefficient representing ratio of IRRs: 1.36, 95% CI: 1.25, 1.48) when preceded by two dry rather than average winters. Incidence in arid lower San Joaquin Valley counties was most sensitive to winter precipitation fluctuations, while incidence in wetter coastal counties was most sensitive to summer temperature fluctuations.

**Conclusions:** In California, wet winters along with hot summers, particularly those following previous dry years, increased risk of coccidioidomycosis in California. Drought conditions may suppress incidence, then amplify incidence in subsequent years. With anticipated increasing frequency of drought in California, continued expansion of incidence, particularly in wetter, coastal regions, is expected.

## Introduction

Coccidioidomycosis is a major cause of community-acquired pneumonia in the southwestern U.S. [1–3]. Infection can lead to a primarily respiratory illness that can last months and may progress to a chronic state in 5-10% of individuals, that can last years or be lifelong [1, 4]. Infection occurs through inhalation of spores of the soil-dwelling fungus belonging to the *Coccidioides* genus, that can become airborne through wind erosion or soil disturbance [1]. The dominant species of *Coccidioides* varies geographically, with *C. immitis* most dominant in California and *C. posadasii* in Arizona and other parts of the southwest. Coccidioidomycosis has seen dramatic increases in incidence and an expansion in geographic range over the past two decades. In California, state-wide age-adjusted incidence rates of coccidioidomycosis increased nearly 800% from 2000 to 2018, and over 300% from 2014 to 2018 [5]. The highest disease burden in California occurs in the southern San Joaquin Valley, but the largest proportional increases in incidence rates are seen among counties that lie outside this hot and arid region where cases were historically concentrated [3, 5, 6]. For instance, incidence in counties within the northern San Joaquin Valley increased by over 1,500% between 2000 and 2018, and incidence in the counties within the central coast increased 800% between 2014 and 2018 [5]. These regions are not uniformly hot and arid, with some exhibiting average precipitation twice that of the historically endemic southern San Joaquin Valley [7]. Changing climatic factors that influence the distribution of suitable *Coccidioides* habitat may play a major role in the recent rise of coccidioidomycosis in California.

Among the most concerning climatic changes recently observed in California is the increase in drought frequency and severity, a trend which may continue under anthropogenic warming [8]. A drought is defined as a period of anomalously dry conditions that results in water-related problems, and can be classified into types [9]. California experienced one of its most severe droughts in recorded history between May 2012 and October 2015, receiving less precipitation in 2013 than in any previous calendar year since records began [8, 10]. The drought caused by anomalously low precipitation during 2012-2015 was exacerbated by record high temperatures [11, 12].

The 2012-2015 drought was preceded by a less severe drought spanning March 2007 to November 2009 [13]. While public records show that statewide coccidioidomycosis incidence was lowest during drought and highest in years immediately following drought [14], the change in coccidioidomycosis incidence attributable to drought has yet to be estimated formally.

Droughts result from anomalously low precipitation and may be exacerbated by high temperatures [11, 12]. Precipitation and temperature are two climatic factors that covary with the geographic range of *Coccidioides* spp. in soil [15]. In California, precipitation and temperature are highly seasonal, with wet and cool winters, and dry and warm summers. Coccidioidomycosis incidence also tends to be seasonal, with incidence rising from spring to a peak in late fall. Periods of precipitation facilitate *Coccidioides* hyphal growth and sporulation [16, 17], and hot and dry periods cause the hyphae to autolyze, liberating infectious, heat-tolerant spores termed ‘arthroconidia’, and permitting dispersal of spores from desiccated soils via wind erosion or soil disturbance [18–22]. While prior studies generally support that the alternating wet and dry periods enhance transmission [19, 21, 23, 24], specific details concerning these wet and dry periods—such as the timing and duration for which they are associated with amplified risk, and the magnitude of this effect—are less clear, and results have been found to vary by model structures, geographic foci, and approaches to disaggregate seasonal trends and account for lagged effects of climate [22, 25–27]. Divergence in results of prior work may stem from an underlying nonlinear relationship between coccidioidomycosis incidence and ambient temperature and precipitation, which would limit inference made from traditional linear models and prevent generalization of results to geographies with differing climates. For instance, while a wet period may support hyphal growth, *Coccidioides* is typically found in areas with <600 mm of annual precipitation, suggesting that excess moisture may limit *Coccidioides* presence [15].

Moreover, sequences of antecedent precipitation and temperature extending not only over the year prior to infection, but over several years, may regulate coccidioidomycosis transmission. Previous studies have reported an effect of seasonal precipitation on transmission of *C. posadasii* in Arizona delayed by as much as 2-3 years [19, 20, 27], but none have examined these delayed effects for *C. immitis* transmission in California, nor whether precipitation in prior years modifies the influence of more recent precipitation effects in the months leading up to transmission.

In this study, we establish a comprehensive understanding of the complex and nonlinear relationships between temperature, precipitation and coccidioidomycosis incidence, examining how these relationships vary across different time periods and geographic areas, and the degree to which the effects of intra-annual climatic factors are modified by inter-annual climatic factors. We estimate the causal effect of the California droughts of 2007-2009 and 2012-2015 on coccidioidomycosis incidence across geographic areas, applying a non-parametric substitution-estimator (G-computation [28]) approach to simultaneously describe space-varying, delayed, and nonlinear effects. By filling critical knowledge gaps about the region-specific roles of seasonal climate and drought on coccidioidomycosis, we strengthen our understanding of why California has observed substantial increase and geographic expansion of incidence over the past two decades.

## Methods

### Data

We obtained California Department of Public Health (CDPH) reportable disease surveillance data on confirmed coccidioidomycosis cases reported among California residents with estimated date of disease onset from April 1, 2000 – March 31, 2020. Diagnosed coccidioidomycosis cases are required to be reported by health care providers and laboratories to local health departments and then the CDPH [2, 29, 30]. Prior to January 1, 2019, a confirmed coccidioidomycosis case in California met both laboratory and clinical criteria of the Council of State and Territorial Epidemiologists (CSTE) 2011 coccidioidomycosis case definition, although case definition compliance varied by local health jurisdiction [31]. Following January 1, 2019, confirmed cases were only required to meet coccidioidomycosis laboratory criteria as defined by CDPH [32]. Per CDPH protocols, a reported coccidioidomycosis case can only be reported once per person, so patients with reactivation of infection were included only for their initial diagnosis. Using an offline geocoding routine in ArcGIS [33], we determined the census tract of each case’s residence based on reported patient address. Where street address could not be identified (11% of patients) we used the centroid of the zip code. Overall, we matched 95% of patients to a location.

Coccidioidomycosis case data was summarized as the total number of cases recorded each month, within each census tract. Environmental and climate data was also summarized as monthly vales at the census-tract level; for each census tract, we calculated the total monthly precipitation, the mean daily average temperature. We averaged time-invariant landscape features including elevation and soil texture across census tract. To maintain comparability of temporal trends with published records, patients were assigned to the month in which the surveillance record indicated estimated disease onset. However, for many patients, this was also the date of specimen collection, which may have occurred weeks following symptom onset. We obtained daily minimum and maximum temperature and daily precipitation from PRISM from 2000– 2020 at 4 km resolution [7]. We obtained information on soil texture (e.g., percent sand, silt, and clay) from POLARIS, a 30-meter probabilistic soil series dataset for the contiguous United States [34]. We obtained information on the fractional coverage of impervious surface from the National Land Cover Database [35], and on elevation from United States Geologic Service (USGS) National Elevation Database [36].

We focused our analyses on geographic areas with notable incidence rates and case counts. Because the Sierra Nevada and San Emigdio-Tehachapi mountains produce strong gradients in temperature and precipitation within the boundaries of some counties with high incidence, we split the counties of Kern, Fresno, Madera, and Tulare into eastern and western regions, and Los Angeles into northern and southern regions, along a 500-meter elevation isocline. We defined the study region to include counties or sub-counties where cumulative cases exceeded 500 cases over the study period and mean annual incidence rate exceeded 10 cases per 100,000 population (Figure 1A). Fourteen counties or sub-counties were included in the final analysis (Figure 1; Table 1), which we hereinafter refer to as “ counties”.

**Figure 1.**
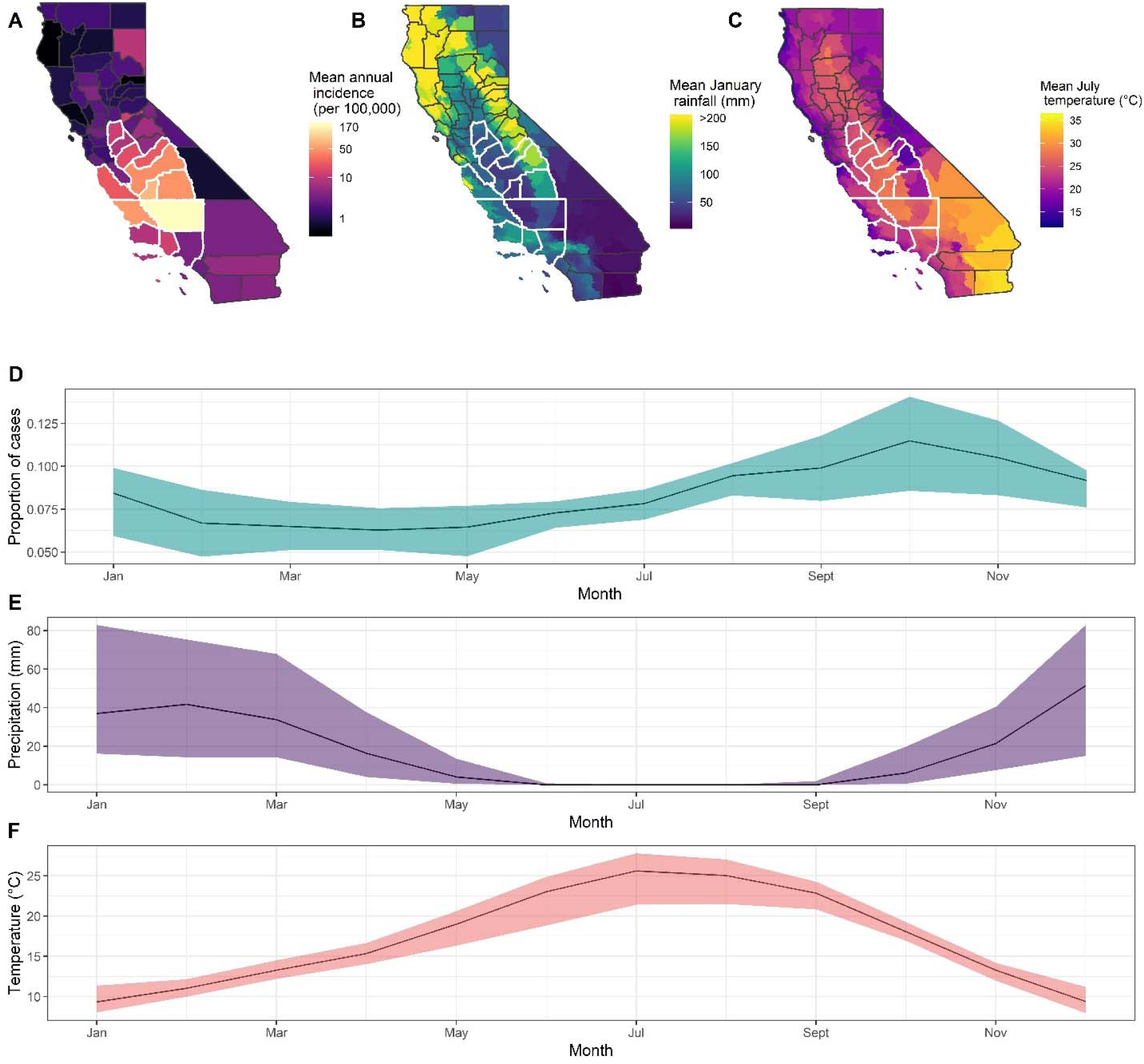
Average annual incidence of coccidioidomycosis (A) between 2000 and 2020. Mean total monthly precipitation (B) during January between 2000–2020. Average daily temperature (C) during July between 2000–2020. Counties outlined in white were included in analyses. Mean (dark line) and interquartile range (shaded area) of the proportion of annual cases (D) with an estimated date of onset per month, between 2000 and 2020 in the study region. Median (dark line) and interquartile range (shaded area) of precipitation (E), in millimeters, per month, between 2000 and 2020 in the study region. Median (dark line) and interquartile range (shaded area) of temperature (F), in degrees Celsius, per month, between 2000 and 2020 in the study region.

**Table 1.** Excess cases associated with California droughts spanning 2007-2009 and 2012-2015. ‘Expected’ represents expected incident cases if below average precipitation and hotter than average temperatures during the drought period had been deterministically set to average levels. ‘Difference’ represents difference between observed incidence and expected incidence, here interpreted as the incident cases attributable to the drought.

|  | May 2012 – October 2015 drought |  |  |  |  |  | March 2007 – November 2009 drought |  |  |  |  |  |
| --- | --- | --- | --- | --- | --- | --- | --- | --- | --- | --- | --- | --- |
| County | During drought |  | 2-yrs following |  | Total |  | During drought |  | 2-yrs following |  | Total |  |
|  | Expected | Difference | Expected | Difference | Expected | Difference | Expected | Difference | Expected | Difference | Expected | Difference |
| Western Kern | 5,355 | -1,965 | 5,949 | 446 | 11,304 | -1,608 | 2,829 | -1,145 | 4,511 | 274 | 7,341 | -960 |
| Western Fresno | 1,638 | -115 | 2,054 | 674 | 3,692 | 591 | 1,203 | 71 | 1,326 | 345 | ,2529 | 434 |
| Western Tulare | 808 | 92 | 924 | 315 | 1,732 | 430 | 588 | 45 | 439 | 106 | 1,027 | 167 |
| San Luis Obispo | 383 | -68 | 811 | 301 | 1,195 | 257 | 274 | -44 | 379 | 144 | 653 | 114 |
| Ventura | 496 | -8 | 627 | -19 | 1,123 | -17 | 189 | -3 | 143 | 2 | 332 | 2 |
| Kings | 458 | -63 | 509 | 132 | 966 | 77 | 295 | 2 | 410 | 102 | 705 | 111 |
| Monterey | 308 | -33 | 642 | 205 | 950 | 181 | 122 | -9 | 174 | 67 | 296 | 60 |
| San Joaquin | 376 | 95 | 556 | 181 | 932 | 291 | 165 | 34 | 192 | 93 | 357 | 132 |
| Northern Los Angeles | 419 | -48 | 427 | 7 | 846 | -32 | 190 | -35 | 222 | 67 | 412 | 48 |
| Eastern Kern | 370 | -48 | 324 | 42 | 694 | -7 | 198 | -33 | 245 | 52 | 443 | 18 |
| Merced | 266 | -7 | 219 | 55 | 485 | 48 | 72 | 0 | 119 | 27 | 191 | 26 |
| Stanislaus | 194 | -25 | 252 | 41 | 446 | 14 | 77 | -7 | 132 | 42 | 209 | 33 |
| Santa Barbara | 151 | -13 | 234 | 51 | 384 | 43 | 69 | -3 | 78 | 17 | 147 | 18 |
| West Madera | 146 | 12 | 127 | 30 | 272 | 44 | 78 | 2 | 77 | 20 | 155 | 22 |
| **Total** | 11,367 | -2,192 | 13,654 | 2,461 | 25,021 | 313 | 6,349 | -1,126 | 8,446 | 1,358 | 14,795 | 228 |

### Distributed lag nonlinear regression models to estimate associations between climatic factors and incidence

Associations between coccidioidomycosis incidence and temperature and precipitation were estimated using a meta-analytic approach for estimating nonlinear, delayed effects across spatial locations [37–39]. We restricted this stage of our analysis to patients with an estimated disease onset (as described by the surveillance record) between September through November—when most cases are typically reported in in California. As the effect of seasonal and lagged climatic factors may vary by season of disease onset [9, 13], doing so enables more clear identification of the effect of climate in distinct seasons while improving comparability of our results with prior results. Moreover, incidence during September through November is strongly correlated with total incidence in a transmission year.

We first estimated county-specific associations of lagged monthly average temperature and total precipitation and monthly incidence using distributed-lag generalized additive models [38]. Full model details are included in the Supplement. We used monthly cases per census tract as the outcome variable and the log of each census tract’s population as an offset term so that model coefficients reflected the log incidence rate ratio. The primary exposure variables were lagged total precipitation and mean temperature. These were modeled with natural cubic spline functions of smoothed three-month averages, with lags spanning 1 to 36 months prior to estimated date of onset. We also included a natural cubic spline for soil type (percent sand) and year. To determine the location of knots for the cubic spline, we systematically varied the location of internal knots placed at precipitation or temperatures corresponding to average percentiles across counties [38], selecting the model that minimized the sum of Q-AIC across all counties, where the Q-AIC is a modification of the Akaike information criterion (AIC) for quasi-likelihood models [40]. We identified the 25^th^ and 75^th^ percentiles of precipitation or temperature for the lagged months included in a 12-month cycle (e.g., a lag of one month corresponds to August - October). We calculated the incidence rate ratio per interquartile range (IQR) increase in temperature or precipitation as the incidence rate at the 75^th^ percentile compared to the incidence rate at the 25^th^ percentile, keeping other factors fixed.

Then, we used a fixed-effects meta-analysis to pool estimates of county-specific incidence rate ratios [38]. We examined the overall shape of the spline relating precipitation and temperature to incidence at each of the 36 monthly lags, and calculated the pooled IRR associated with an increase of one IQR in precipitation or temperature over each lag. We assessed factors explaining heterogeneity in the temperature-incidence and precipitation-incidence relationships across counties by extending the fixed-effects meta-analysis to include meta-predictors of county-level information (e.g., mean county total precipitation, mean county temperature). We determined the significance of the meta-predictors using Wald tests [38] and plotted separate exposure-response relationships across our set of significant meta-predictor values.

To examine potential modification of the effect of the wet winter period—the strongest predictor of fall coccidioidomycosis incidence as determined by the fixed-effects meta-analysis—by antecedent conditions, we created a nonlinear interaction term between total monthly winter precipitation per census tract and antecedent conditions by multiplying the basis function for the cubic spline on 9-month lagged precipitation (i.e. recent winter precipitation) by a binary indicator for whether or not the census tract had a drier than average winter in the two years prior to estimated date of disease onset, three years prior to date of onset, or both. The target parameter, the exponentiated coefficient on the modelled interaction term, is thus expressed as the ratio of the IRR for an IQR increase in total monthly winter precipitation following a dry year to the IRR for winter precipitation following a wet year. All statistical analyses were conducted in R, version 3.3.1 (R Foundation for Statistical Computing), using the splines and dlnm package for fitting distributed lag generalized additive models and the mvmeta package for performing fixed-effect meta-regression modeling [38, 41].

### Ensemble model to estimate changes in incidence attributable to drought in California

We estimated cases attributable to—or averted because of—major droughts in California between April 1, 2000 and March 31, 2020 using an ensemble modelling approach that predicts incidence under counterfactual scenarios corresponding to the presence or absence of drought [28, 42]. For this stage of the analysis, we examined incidence throughout the entire year, rather than restricting incident cases to September through November. For each county, we modelled monthly cases at the census tract-level using parametric and non-parametric prediction algorithms (see Supplement).

These included a set of generalized linear models with increasing complexity with respect to variables and interaction terms; generalized additive models; and Random Forest [43]. Model predictors varied by candidate algorithm, but could include: season; year; soil texture; elevation; percent impervious surface; total lagged monthly precipitation; and lagged mean temperature. We calculated the sum of squared errors for each algorithm using leave-out-one-year cross-validation whereby the model was fit for all but one year of the time period, and then used to predict the out-of-sample cases. We then generated an ensemble prediction by generating a weighted average of candidate model predictions where the weights were derived using non-negative least squares and were inversely associated with their out-of-sample prediction error.

Drought can be classified in different ways. Here, we examine agricultural drought, which is defined by lack of soil moisture [9], and thus thought to be most relevant for *Coccidioides*. Soil moisture varies with both precipitation and temperature, both precipitation and temperature are considered to be primary exposures of interest. We used a simple substitution estimator (G-computation [28]) to calculate the expected incident cases in census tract *i* in month *t* with covariates, *W_i,t_*, as observed and the primary exposure, *A_i,t_*, set to either observed or counterfactual values for lagged rainfall and temperature (equation 1). For counterfactual, “ no-drought” scenario, we deterministically set any monthly average temperature higher than the historical average and any total monthly precipitation below the historical average to their monthly county-level means during the two droughts (March 2007 - November 2009; May 2012 - October 2015). We summed across specific time periods and across all census tracts in a county to estimate the number of expected cases in a county over a time period (equation 1).

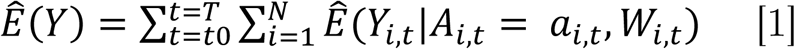

The incident cases attributable to—or averted by—the drought were estimated as the difference between predicted cases under the observed conditions, 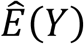, and those predicted under the counterfactual “ average climate” scenario, 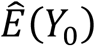(equation 2).

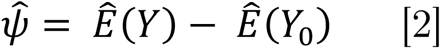

Because antecedent conditions as far back as three years may carry influence, we examined the attributable incidence separately for the 2007-2009 and 2012-2015 droughts in the two years following the end of each drought. Because seasonality of coccidioidomycosis in California is such that incidence is lowest in March-April, with peaks occurring in the fall, we considered the change in incident cases “ during drought” to include the period starting at the onset of the drought and extending until the end of the transmission season following the drought (e.g., March 1, 2007 - March 31, 2010; May 1, 2012 - March 31, 2016). The two years post drought encompassed the full epidemiological seasons following the drought (e.g., April 1, 2010 – March 31, 2012; April 1, 2016 – March 31, 2018).

## Results

### Descriptive analyses

Between April 1, 2000 and March 31, 2020, there were 81,448 reported coccidioidomycosis cases throughout California reported via California Department of Public Health (CDPH) reportable disease surveillance. There were 62,002 (76.1%) cases among residents of the examined counties (Figure 1A), of which 33% of patients had an estimated onset in September – November, 25% in December – February, 24% in June – August, and 18% in March – May (Figure 1D). The counties with the highest average annual incidence rate were Kern (170 cases per 100,000 individuals), Kings (104 cases per 100,000), and San Luis Obispo (43 cases per 100,000) (Figure 1A).

Among the counties analyzed, precipitation and temperature exhibited strong seasonal patterns. From summer lows, precipitation increased starting in September and October and typically peaked between December and February (Figure 1E), exhibiting substantial spatial variation with the lowest mean annual precipitation observed among the southern San Joaquin Valley counties, and the highest in the north and west (Figure 1B). Air temperature typically peaked in July and was lowest in December and January (Figure 1C; 1F).

### Association between precipitation, temperature and fall (September – November) incidence across time and space

#### Effect of recent precipitation and temperature (1-4 months lag)

When analyzing data on cases with estimated onset from September through November, interquartile range (IQR) increases in precipitation in the one to four months prior to the estimated date of disease onset (i.e., during the typically dry summer and early fall), were negatively associated with fall incidence (IRRs by lag: 1 month prior: 0.87, 95% CI: 0.80, 0.94; 2 prior: 0.89, 95% CI: 0.85, 0.94; 3 prior: 0.95, 95% CI: 0.92, 0.98; 4 prior: 0.87, 95% CI: 0.81, 0.94; Figure 2A; Table S1). That is, increasing precipitation in the month prior to estimated disease onset from the 25^th^ percentile to the 75^th^ percentile was associated with a 13% reduction in incidence rates in September – November. The shape of the relationship between recent precipitation and incidence (Figure 3A) indicated a suppression of incidence as precipitation increased from 0 mm (no summer precipitation) to 6 mm, with subsequent increases in precipitation exhibiting lower marginal effect. Exposure-response relationships between precipitation and incidence at all lags are shown in Figure S1.

**Figure 2.**
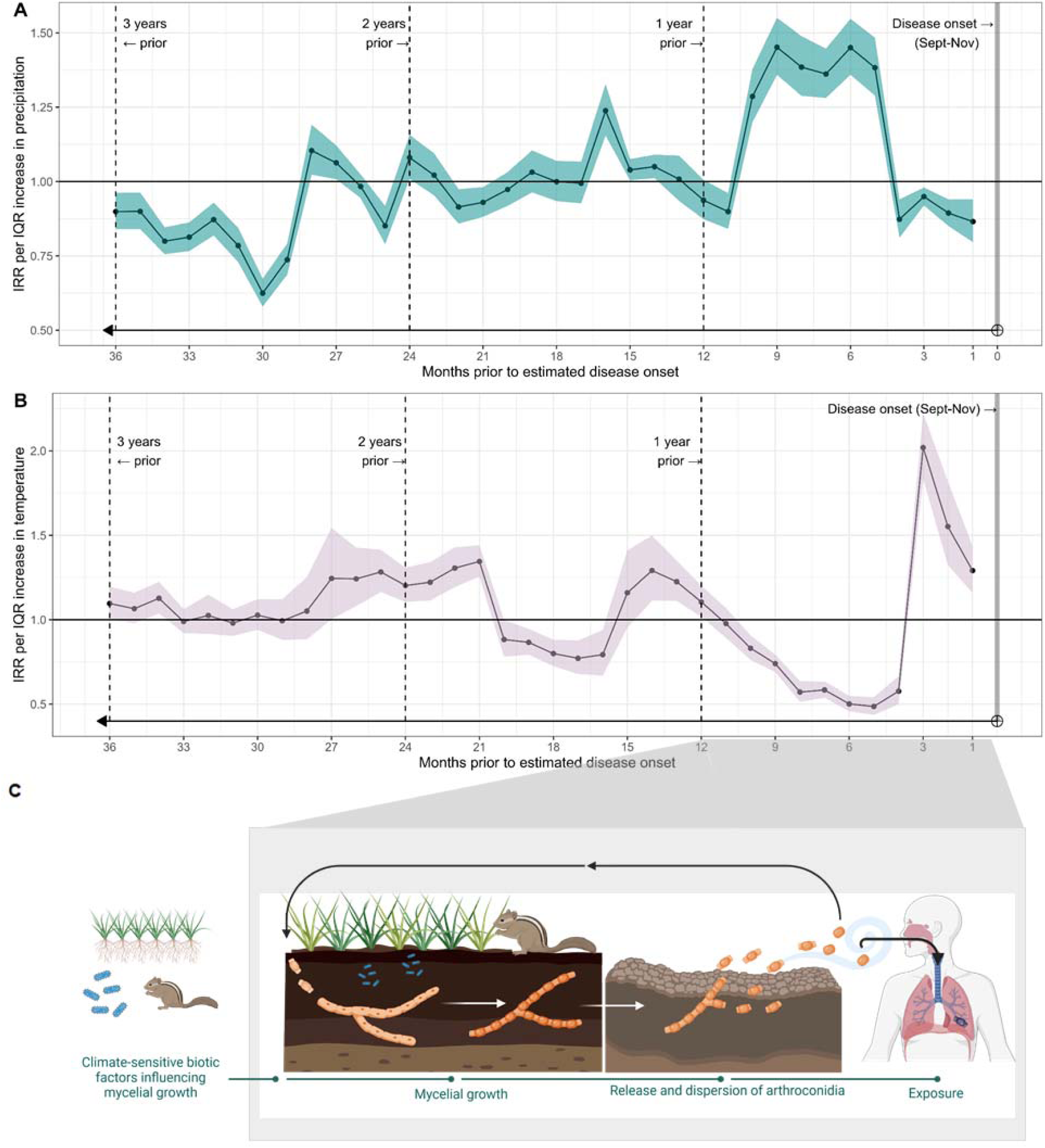
Results of distributed lag, generalized additive model testing the association between fall (September – November) incidence rates and lagged meteorological variables. Incidence rate ratios (IRRs) express the effect of an IQR increase in precipitation (A) or temperature (B) in months prior to the estimated date of disease onset, with confidence intervals shown by shading. The horizontal line at one indicates null association (IRR=1). Panel C displays the saprobic lifecycle of *Coccidioides* and maps the hypothesized grow and blow cycle to the intra-annual wet-dry patterns. Inter-annual influences affecting mycelial growth might include biota, small mammals, and soil dwelling microbial competitors (in blue). These factors may be influenced by climate across inter-annual time scales.

**Figure 3.**
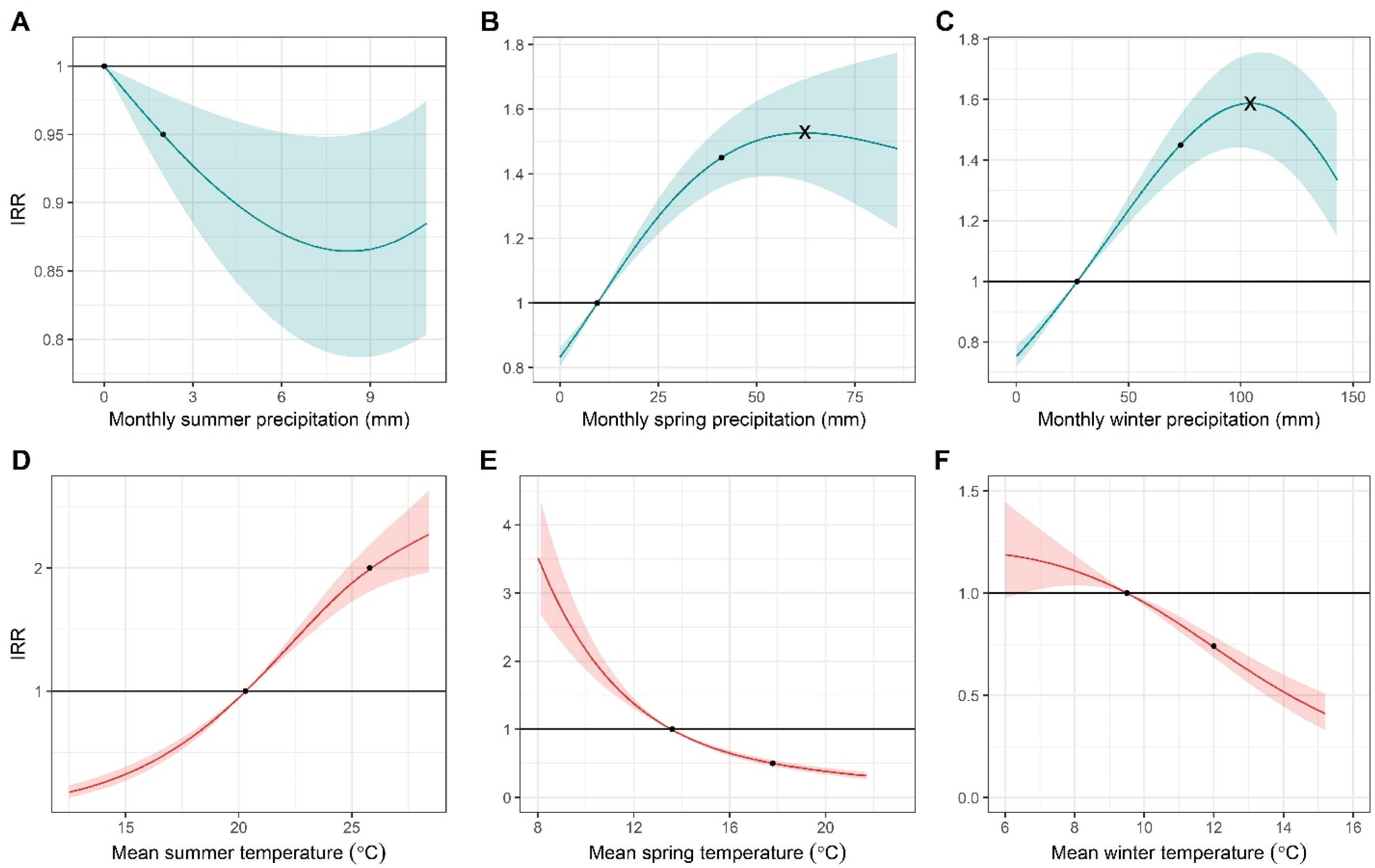
Pooled incidence rate ratios (IRR; solid lines) corresponding to the effect on fall (Sept – Nov) incidence of changing precipitation (A-C) and temperature (D-F) at lags corresponding to summer (A, D), spring (B, E), and winter (C, F) conditions from a reference level to the value shown. The reference level for the IRR is the 25^th^ percentile for the study region (for which IRR=1), such that the solid curved line indicates the incidence rate ratio for fall incidence at a given temperature or precipitation value compared to the incidence rate at the 25^th^ percentile condition for the study region in that time period. Shaded regions reflect the 95% confidence interval. Precipitation corresponding to the maximum IRR is indicated by an ‘x’ in panels B and C. Black circle markers indicate the IRR corresponding to the 25^th^ and the 75^th^ percentile of the exposure values.

Average daily mean temperature in the one to three months prior to estimated disease onset, during the typically hot summer months, was positively associated with coccidioidomycosis incidence (IRRs by lag: 1 month prior: 1.29, 95% CI: 1.16, 1.44; 2 prior: 1.55, 95% CI: 1.32, 1.82; 3 prior: 2.02, 95% CI: 1.84, 2.22; Figure 2B, Table S1). The exposure-response relationship at a 3-month lag (Figure 3D) showed that fall incidence increased with increasing summer temperature monotonically, with no apparent maximum beyond which temperatures are too hot. Exposure-response relationships for all lags are shown in Figure S2.

#### Effect of lagged precipitation and temperature (5-10 months lag)

Positive, significant pooled associations were detected between precipitation lagged 5-10 months and coccidioidomycosis incidence during September through November. The association peaked for precipitation in the winter prior to estimated date of disease onset (i.e., precipitation lagged 9 months; Figure 2A; Table S1). An increase of total monthly winter precipitation from the 25^th^ percentile (27.1 mm) to the 75^th^ percentile (73.2 mm) in the 9 months preceding disease onset was associated with a 45% (IRR:1.45, 95% CI: 1.36, 1.55) increase in coccidioidomycosis incidence in the fall. Pooled exposure–response relationships for both winter and spring precipitation showed a unimodal response whereby an increase in coccidioidomycosis incidence was observed with incremental increases in precipitation until an optimal value was achieved (around 40-65 mm during spring, 80-105 mm during winter; Figure 3B, 3C), after which additional precipitation was associated with lower incidence as compared to the optimal.

Higher temperatures in the winter and spring prior to estimated date of disease onset (i.e., 5-10 months prior) were associated with suppressed incidence. In pooled analyses, an increase of one IQR in average monthly temperature in the winter prior to estimated date of disease onset (from 9.5°C to 12°C) was associated with a 26% (IRR: 0.74, 95% CI: 0.69%, 79%) decrease in incidence rates in the fall. Fall incidence increased as winter temperatures (i.e., 9-month lagged temperatures) dropped, until around 8°C, below which the effect of temperature was uncertain due to sparse data (Figure 3F).

#### Antecedent precipitation and temperature (>12 months lag)

Antecedent precipitation and temperature occurring in the two to three years prior to estimated disease onset were associated with coccidioidomycosis incidence (Figures 2B and 2C). While total monthly precipitation in the winter immediately prior to estimated date of disease onset was positively associated with incidence, precipitation in the winters two and three years prior to estimated date of disease onset were negatively associated with incidence (Figure 2A). For temperature, warmer summers and cooler springs occurring two to three years prior to disease onset were associated with higher incidence, with a dampening of this association seen as the lag increased (Figure 2B).

Antecedent conditions were important effect modifiers of the effect of more recent meteorological conditions. While precipitation in the most recent winter was the strongest predictor of coccidioidomycosis incidence when compared to other lags, precipitation in the winters 2-3 years prior was found to be a significant effect modifier of this relationship, with antecedent dry conditions amplifying the effect of wet winters on incidence. Specifically, when following a year with low winter precipitation (i.e., a year with winter precipitation falling below the 50^th^ percentile), an IQR increase in current year winter precipitation was associated with an IRR 1.19 (95% CI: 1.10, 1.30) times larger than the IRR when the same IQR increase in current year winter precipitation was experienced following a year with high winter precipitation (i.e., a year with winter precipitation above the 50^th^ percentile). When following two years of below median precipitation, the effect of a one IQR increase in current year winter precipitation was 1.36 (95% CI: 1.25, 1.48) times the effect of the same increase in winter precipitation following two years in which precipitation for both years was not drier than average. Put another way, the incident cases attributable to precipitation in a wet winter prior to estimated date of disease onset was highest for wet winters that followed two consecutive dry years.

#### Heterogeneity in effects by precipitation and temperature gradients

In multivariate meta-regression models, we found that the typical winter precipitation (i.e., the median winter precipitation) in counties explained a significant amount of heterogeneity in county-specific IRRs representing the effect of winter precipitation on incidence (Figure 4A). The effect of a one IQR increase in winter precipitation (from 27 mm to 73 mm) was most pronounced among counties where the median monthly winter precipitation was low. For instance, in western Kern, which experiences only 22.7 mm of total precipitation in a typical (50^th^ percentile) winter month, an increase from 27 to 73 mm of precipitation was associated with an IRR of 1.67 (95% CI: 1.42, 1.96). While the pooled exposure-response relationship was strongly nonlinear, very dry regions, such as western Kern county, rarely if ever exhibit winter precipitation high enough to see the maximum amplification of incidence attributable to precipitation (indicated by the ‘x’ in Figure 3C). As the median precipitation per county increased, the effect of precipitation on incidence attenuated (Figure 4A). Precipitation in the wettest counties, such as Monterey, which typically receives 70 mm of precipitation in a single winter month, had a non-significant negative effect on incidence. In contrast, there was little heterogeneity in the county-specific effects of summer precipitation on incidence, as most counties examined experienced little to no precipitation during these months. Thus, spatial variation in winter precipitation drives heterogeneity in the delayed effect of rainfall on coccidioidomycosis incidence, with dry counties most sensitive to fluctuations in precipitation.

**Figure 4.**
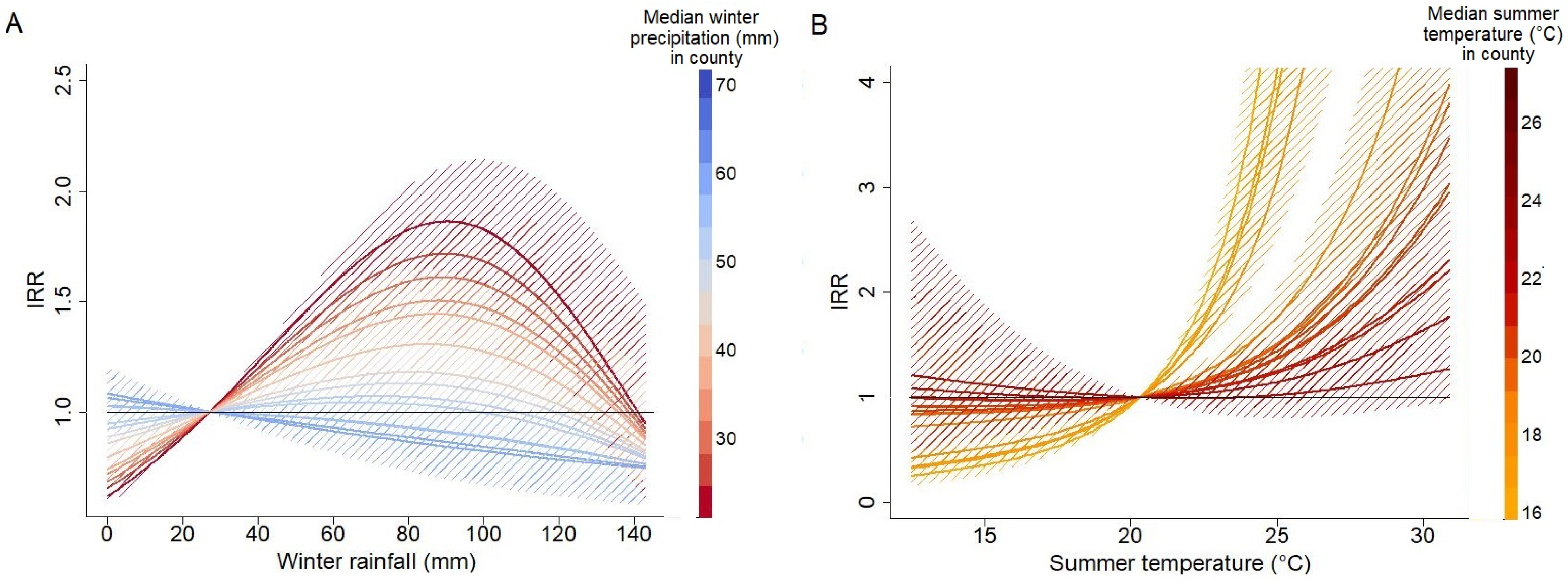
Increases in winter precipitation or summer temperature had the greatest effect in regions where rain was scarce or temperatures were low, respectively. (A) Estimated exposure-response relationships expressed as incidence rate ratios (IRR; colored lines) corresponding to the effect of changing winter precipitation and (B) summer temperature from a reference level to the value shown. The reference level for the IRR is the 25^th^ percentile for the study region (for which IRR=1). Each line indicates an exposure-response relationship expressed as the incidence rate ratio for a given temperature or precipitation value compared to the incidence rate at the 25^th^ percentile condition for a given county, based on the county’s median winter precipitation (A) or median mean summer temperature (B). Dashed regions around the solid lines indicate 95% confidence intervals.

For temperature, multivariate meta-regression results likewise suggested that typical summer temperature (i.e., the median summer temperature) in counties explained a significant amount of heterogeneity in county-specific effects of summer temperature on incidence (Figure 4B). The effect of a one IQR increase in summer temperature (from 20.3°C to 25.8°C) was most pronounced among counties where the median summer temperature was coolest. For instance, in Monterey, which experienced a mean monthly temperature of 16.2°C in a typical summer month, an increase from 20.3°C to 25.8°C in temperature was associated with an IRR of 12.7 (95% CI: 3.07, 53.3). An increase in mean summer temperature in the hottest 4 of 14 counties, such as western Kern, which typically experiences a mean month temperature of 27.0°C, was non-significant (IRR in Kern: 0.79, 95% CI: 0.48, 1.30). There was little heterogeneity in the county-specific effects of winter temperature on incidence. In summary, spatial variation in summer temperature drives heterogeneity in the delayed effect of temperature on coccidioidomycosis incidence, with cooler counties most sensitive to fluctuations in temperature (Figure 4B).

### Coccidioidomycosis incidence attributable to drought in California (ensemble models)

Ensemble models were able to explain over 90% of the variation in coccidioidomycosis incidence in the study region. Counterfactual model predictions aggregated across the study period (Figure 5A; with county-specific model fits shown in Figures S5-17) revealed lower than expected transmission during periods of drought, with higher than expected transmission across the two years following (Table 1). Higher excess incidence attributable to drought were seen following the more severe 2012-2015 drought (Figure 5C) as compared to the 2007-2009 drought (Figure 5B).

**Figure 5.**
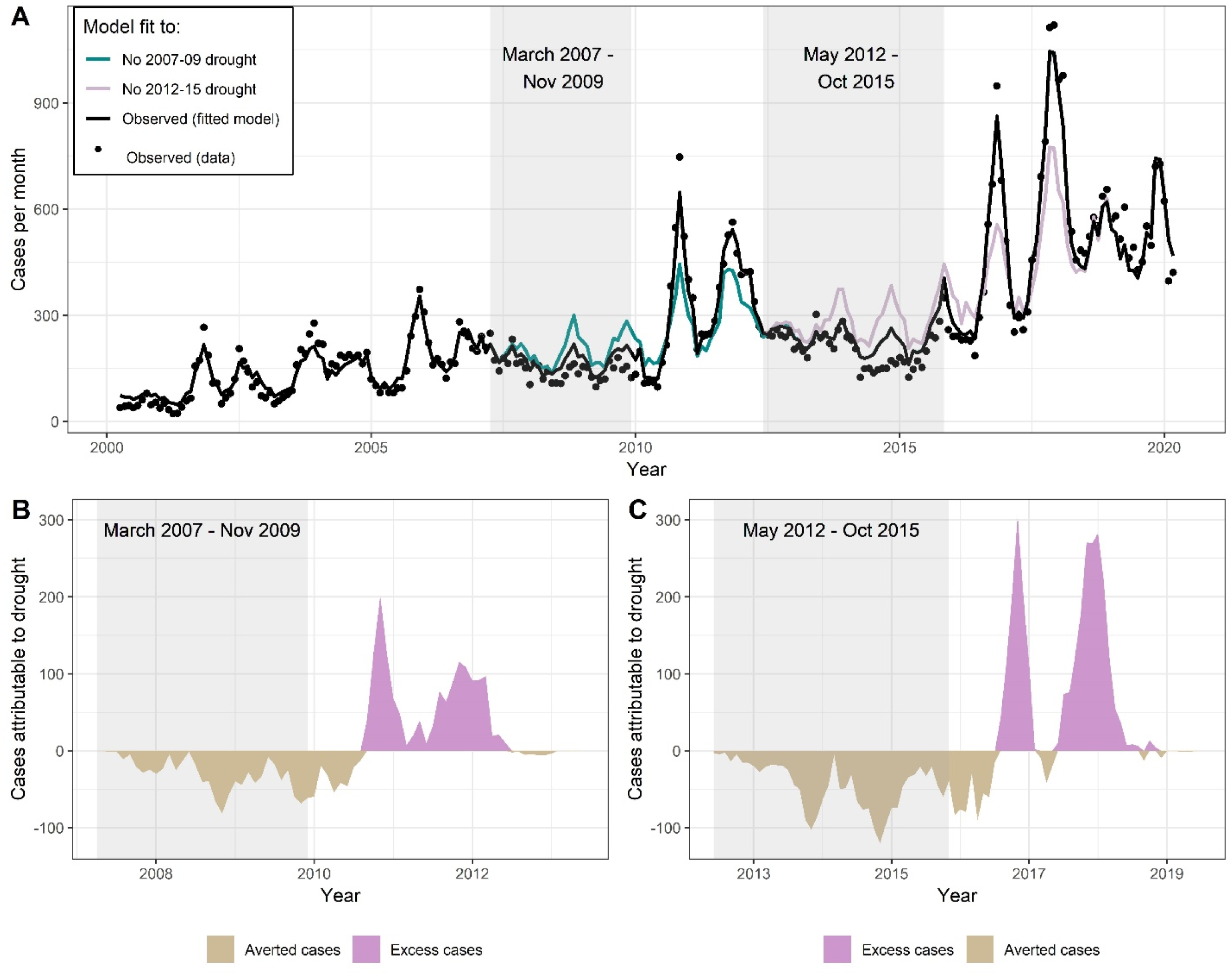
Droughts were associated with reduced incidence during the drought, and excess incidence following the drought. (A) Observed incidence (black dots) by month within the study region. Black line is the model fit under the observed environmental conditions. Color lines represent the expected incidence under the counterfactual intervention if the 2007-09 drought did not occur (cyan) or the 2012-15 drought did not occur (pink). Counterfactual scenarios were generated by setting temperatures observed to be higher than historical averages, and precipitation values observed to be below historical averages, deterministically to their average values. Gray boxes indicate the drought period. (B and C) Difference between expected cases and counterfactual cases if the 2007-09 (B) and the 2012-15 (C) droughts had not occurred, respectively.

In 11 of the 14 counties examined, the 2012-2015 drought was associated with a decline of cases below expected during the drought, followed by an increase in cases above expected in the two years following the drought (Table 1; Figure 6 for relative changes, Figure S4 for absolute changes). In all counties except for western Kern, the increase in cases following the drought exceeded the decrease in cases during the drought. In the counties examined, drought was associated with 2,172 fewer cases between May 1, 2012 – March 31, 2016, and 2,460 excess cases between April 1, 2016 –March 31, 2018.

**Figure 6.**
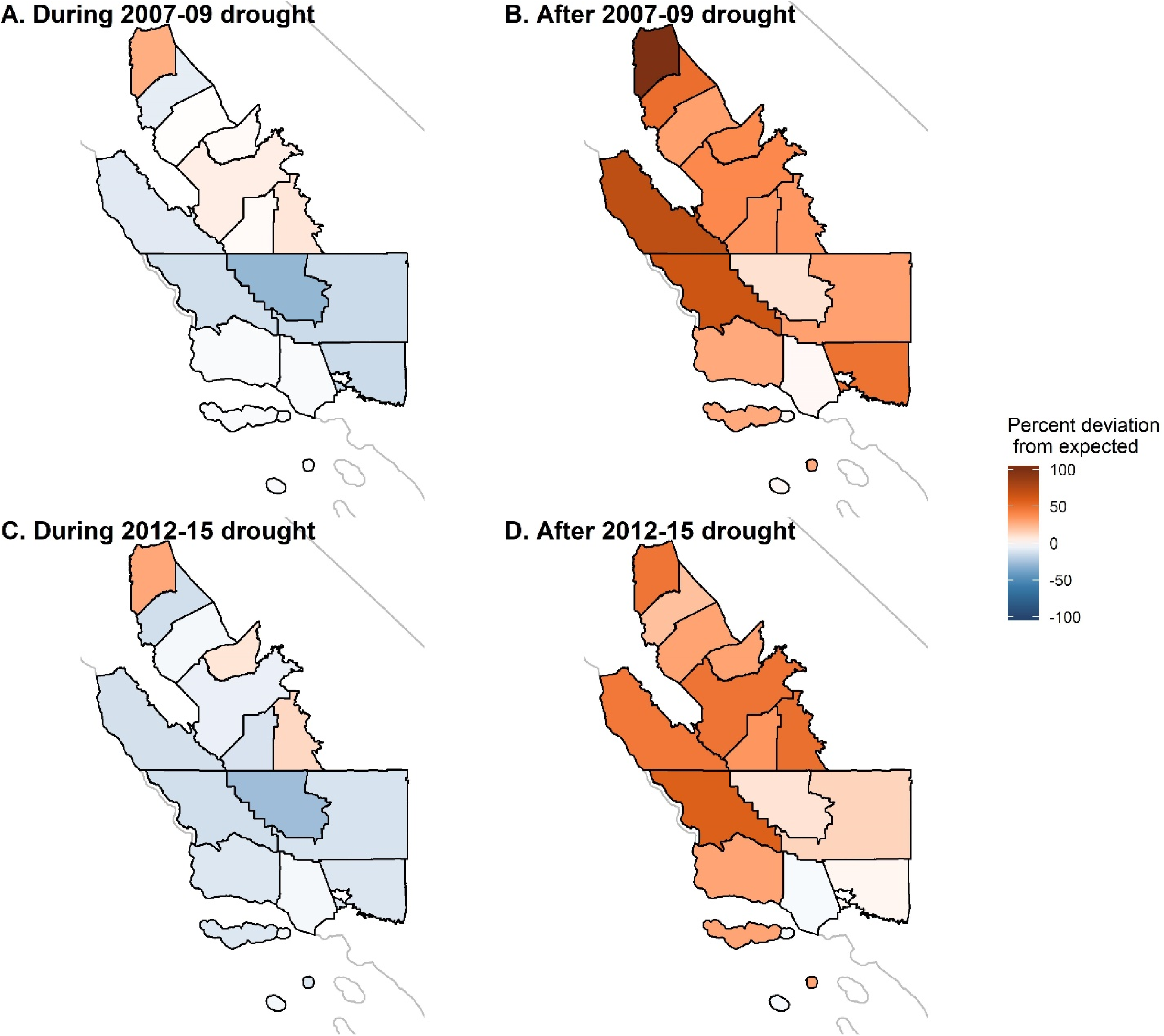
Estimated percent deviation in incident cases compared to the number expected in the absence of drought during (A and C) and in the two years following (B and D) the 2007-09 (A, B) and 2012-15 (C, D) droughts across the 14 counties in the study region. California state outline shown in light gray.

Kern County west of the Sierra-Nevada Mountains has the largest burden of coccidioidomycosis in the state. Over the 47 months spanning May 1, 2012 – March 31, 2016 drought period, 3,390 cases of coccidioidomycosis were reported among residents of western Kern County. Had conditions not been drier or hotter than average (i.e., absent the drought), there would have been an estimated 1,965 more cases of coccidioidomycosis during this period. Thus, drought conditions were associated with an estimated 36.7% reduction from expected (counterfactual) incidence over May 1, 2012 – March 31, 2016 in Kern County. However, in the 24 months following the drought— when Kern County observed 6,394 cases—we estimated that if the drought had not taken place, the county would have seen 445 fewer cases. Thus, drought was associated with a 7.5% increase from expected incidence in the 24 months following the drought in Kern County.

The 2007-2009 drought followed similar patterns as the 2012-2015 drought but was associated with fewer averted cases during the drought period, while being associated with a similar number of excess cases in the two epidemiological years (April - March) that followed. As a result, this drought—which was shorter in duration and experienced lower temperatures than the later 2012-2015 drought—also resulted in a net increase in cases. Across all counties examined, the drought was associated with 1,125 fewer cases between March 1, 2007 – March 31, 2010, and 1,358 excess cases between April 1, 2010 – March 31, 2012 in the study region. The decline in cases during the drought was most prominent in western Kern County and least pronounced in the counties in the northern San Joaquin Valley, while the increase following the drought was most prominent among the coastal counties and those in the northern San Joaquin Valley (Figure 6; Figure S4). An estimated 1,145 cases were averted in western Kern county during the 37 months spanning March 1, 2007 – March 31, 2010, corresponding to a decline of 40.4% from expected. There was an estimated excess of 273 cases during the 24 months following the drought from April 1, 2010 – March 31, 2012 in Kern County, corresponding to an increase of 6.1% from expected.

## Discussion

This work finds that drought temporarily displaces coccidioidomycosis transmission, suppressing cases in years characterized by drought conditions but amplifying cases in years immediately following drought conditions. Across 14 counties examined, we estimated that an excess of 1,358 and 2,461 drought-attributable cases were reported in California following the 2007-2009 and 2012-2015 droughts, respectively, more than offsetting the drought-attributable decline of 1,126 cases during the 2007-2009 drought and 2,192 during 2012-2015 drought. This represents a previously underexplored health consequence of drought in California. Meanwhile, the impact of seasonal variation in climatic factors on coccidioidomycosis transmission, irrespective of drought, underscore this trend: in general, cases increased following wetter than average winters and hotter than average summers. The magnitude of this effect was mediated by the underlying average climate regime of a given region. Overall, our findings motivate inclusion of drought and winter rainfall in incidence projections, anticipation of different effects of drought and precipitation across geographic regions, and intensification of prevention measures in wet years that follow drought.

Our findings can be interpreted in the context of specific lifecycle stages of the fungus, as well as the major hypotheses that link coccidioidomycosis transmission to environmental processes (Figure 2C; Table 2). First, associations detected between recent climatic conditions and coccidioidomycosis incidence likely signal the suppression or amplification of mycelial fragmentation into spores, and spore dispersal, while associations detected between coccidioidomycosis incidence and climatic conditions at longer time lags (e.g., 5 months or more) likely signal mechanisms involving long-term growth of mycelia and survival of spores in the environment (Figure 2C). The “ grow and blow” hypothesis is among the most widely accepted mechanistic theories regarding the influence of climate conditions occurring within the past year on seasonal coccidioidomycosis [19, 21, 23, 24], and other soil-dwelling fungal pathogens may take advantage of this strategy [44]. We support and enhance our understanding of this hypothesis by suggesting that, within an annual cycle, hyphal growth for *C. immitis* may be most important during winter and spring months when there is adequate rainfall to promote growth [17]. Pooled regression estimates show that the strongest effect of precipitation on coccidioidomycosis incidence is observed at a 9-month lag before disease onset in the fall (Figure 2A), corresponding to winter precipitation. Meanwhile, a higher temperature and lower precipitation during summer and fall was associated with increased incidence, suggesting that lysis of hyphae into singular spores and wind dispersion of spores is most important in the summer and fall.

**Table 2.** Summary of findings from generalized additive modeling of lagged precipitation and temperature and coccidioidomycosis incidence in the present study, and their relationship to established hypotheses and specific lifecycle components of *Coccidioides* spp.

| Time period relative to estimated date of disease onset | Hypothesized lifecycle function | Hypothesis in question | Main finding on influence of meteorology | Variation in findings across space | Relationship to drought |
| --- | --- | --- | --- | --- | --- |
| Fall and summer (1-4 months prior to estimated date of disease onset) | Fragmentation of mycelia; dispersal and inhalation of spores via wind erosion | Blow of the ‘Grow and Blow’ hypothesis | High temperature is the driving factor increasing risk. Dry conditions also increase risk. | Most counties in the region examined are dry during fall and summer. Spatial variation in incidence rates is likely driven by spatial variation in temperatures, with some counties several degrees hotter than others, on average. Cooler counties may be heat-limited. | Higher temperatures during drought may increase cases, but the decline of precipitation in spring and winter seen during droughts seems to overcome this effect. |
| Spring and winter (5-10 months prior to estimated date of disease onset) | Growth of mycelia | Grow of the ‘Grow and Blow’ hypothesis | Precipitation is the driving factor increasing risk, with unimodal relationship observed. Cooler temperatures also increase risk. | Spatial variation in spring and winter precipitation is observed. Precipitation in dry counties are moisture-limited, and never exceed the maximum threshold beyond which additional precipitation suppresses risk. Precipitation in wet counties may surpass this threshold. We call these counties “unsaturated” and “oversaturated”. | Low precipitation during drought may be driving the decline in cases seen among most regions during drought. |
| Two to three years prior to estimated date of disease onset | Growth and survival of spores and mycelia; regulation of upstream factors including: other microbial competitors, soil stabilizing vegetation, nutrients such as small mammals and vegetation | Soil sterilization | Antecedent climate modifies the effect of recent climate. The increased risk due to recent winter precipitation is most pronounced following consecutive years of drier than average conditions | Not examined | Incidence following drought is higher than expected. |

**Table 3.** Model choice depended on the research question and associated target parameter.

| **Purpose of analysis** | **Analysis used** | **Target parameter** | **Justification for analysis** |
| --- | --- | --- | --- |
| Estimate the nonlinear relationships between lagged temperature and precipitation and coccidioidomycosis incidence. | Distributed lag generalized additive model at the county-level, with meta-analysis | Incidence Rate Ratio expressing incidence rate of coccidioidomycosis at some temperature or precipitation level to the incidence rate at another level. | Here, the target parameter of interest can be directly expressed from the model coefficients on lagged temperature and precipitation. This approach provides the level of flexibility needed, while permitting Wald-based inference in the form of standard errors. Splines permit the modelling of non-linear relationships. Modelling at the county-level and creating a pooled relationship using meta-analysis permits examination of the factors that explain heterogeneity across space. |
| Estimate how distal climatic factors modify the effect of more recent climatic factors on coccidioidomycosis incidence. | Distributed lag generalized additive model with interaction terms | Model coefficient on the interaction term, reflecting the ratio of incidence rate ratios. | Here the target parameter of interest can be directly expressed from the model coefficients on the interaction term. This approach provides the level of flexibility needed, while permitting Wald-based inference in the form of standard errors. |
| Estimate the number of cases averted by or attributable to droughts in California between 2000 and 2020. | Ensemble prediction model | The absolute difference in cases between observed drought conditions, and counterfactual non-drought conditions, expressed as $E(Y) - E(Y_0)$. | Here, the target parameter of interest is the difference in model predictions, rather than a simple model coefficient. Since we modelled the full time series, a greater level of flexibility in model form was needed to allow for different effects of climatic lags across seasons. This approach provides the most flexibility, while enabling robust predictions. The expense is that model-based inference is limited. |

**Table 4.**
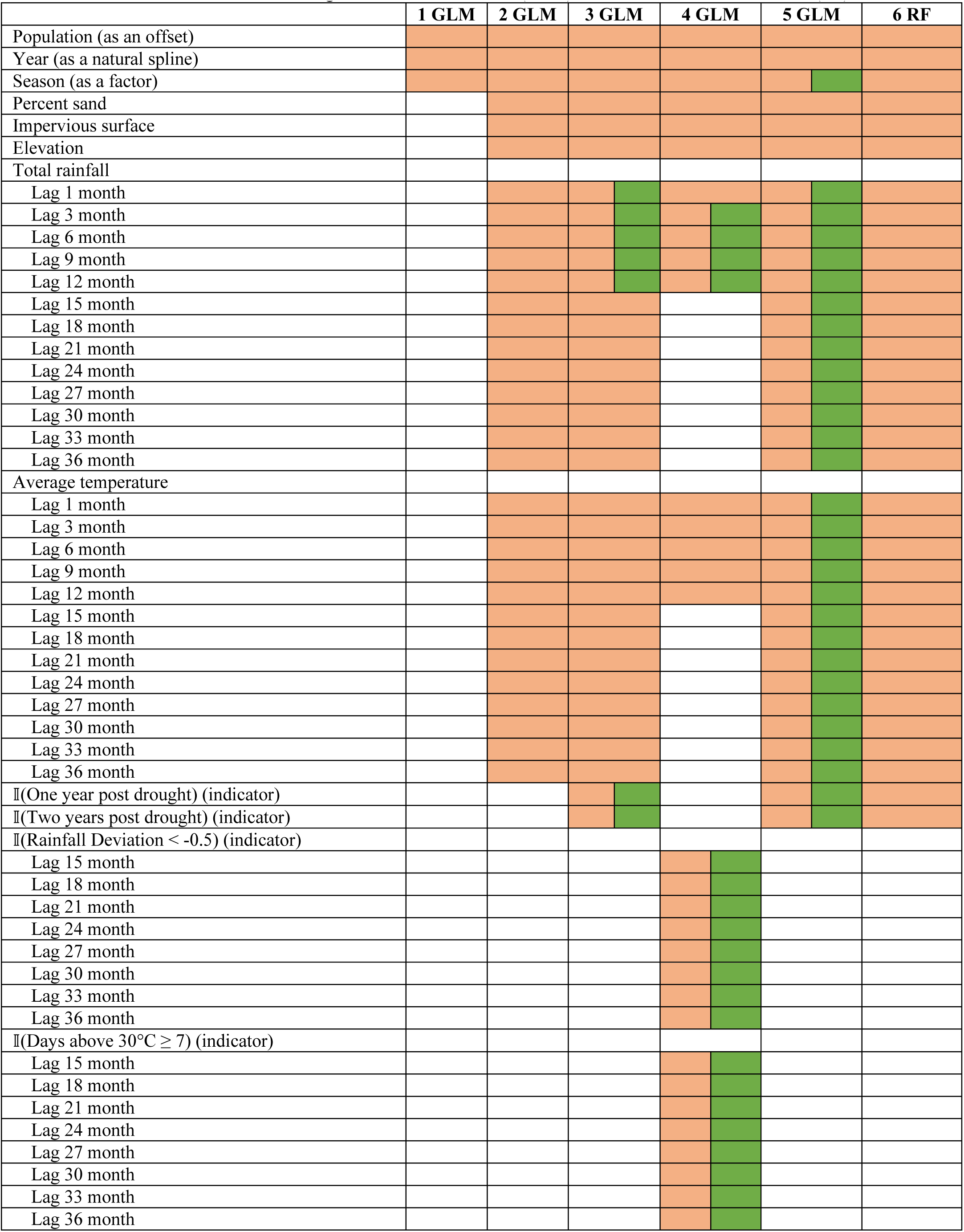
Variables included in the 6 models used in the ensemble model. Orange cells are used as main effects, and green cells were used as interactions. Models 1-5 were generalized linear models (GLM) and model 6 was random forest (RF)

Associations detected between coccidioidomycosis incidence and climatic conditions that occurred more than a year prior may involve the influence of upstream factors on pathogen proliferation, such as nutrient availability and presence of other soil microbes that interact with *C. immitis* (Figure 2C). Both our ensemble model and regression results suggested that an increase in the mycelial growth that occurs during wet periods is induced by prior dry conditions. Wet conditions nine months prior to disease onset yielded substantial increased incidence (Figure 2C), but the effect of this wet period was 36% stronger when it followed a two-year dry period compared to when it did not. There are several hypotheses that may explain these acyclical inter-annual patterns observed over a multi-year time period. First, these findings could lend support to the “ soil sterilization” hypothesis. Laboratory studies suggest that *Coccidioides* spp. are poor competitors for nutrients when compared with certain other soil fungi and bacteria [45, 46], but are resilient and can survive climatological extremes, including low precipitation and intense heat, to which other species may be more susceptible [47].

Thus extreme hot or dry periods may suppress the relative fitness populations of microbial competitors in the soil [19, 24, 48, 49], allowing *Coccidioides* spp. populations to grow uninhibited by competition when more favorable conditions (i.e., rainfall) return. Other plant-microbe interactions may be at play. Genomic studies of *Coccidioides* indicates a lack of plant-metabolizing enzymes, raising the possibility that *Coccidioides* lives mutualistically in soil with microbes that produce exoenzymes that permit *Coccidioides* to access nutrients [50]. Many bacteria may inhibit fungal growth [51]; bacteria such as *Bascillus subtilis* and *Streptomyces* spp. produce antimicrobial compounds that act as antifungal agents [52, 53]. Certain plants exhibit selective forces for the establishment of bacterial and other microbial populations [54].

Other explanations may exist for why antecedent dry conditions influence *Coccidioides* growth once favorable conditions return. An emerging hypothesis is that small mammals are reservoirs, harboring inactive *Coccidioides* granulomas which transform into hyphae following host death [55, 56]. *Coccidioides* has been detected in wild rodents since 1942 [57, 58], and is found in rodent burrows at concentrations over four times greater than that of other soils [46, 55, 59, 60]. Comparative genome analyses demonstrate that the fungus evolved to obtain nutrients from animal substrates more efficiently than plant matter [50]. Rodent death rate is highest during drought [61], which may lead to an accumulation of desiccated keratin in the soil that can be utilized by *Coccidioides* for hyphal growth once more favorable conditions return. In addition, physical and chemical soil properties influenced by precipitation and temperature, including soil pH, salinity, and the presence of flora and fauna, are established over long geological and ecological timescales, and may influence growth of *Coccidioides* spp. [16, 55, 59, 62]. Rodents may influence in other ways. Rodents have been shown to disperse spores of other mycorrhizal fungi [63], and may travel further during drought years to find food. Beyond biological hypotheses, human behavior is an understudied aspect of disease transmission [64]. Human adaptive behaviors to drought, like reduced in agricultural intensity, may play a role in reducing exposure to pathogens [65].

In examining how the underlying temperature and precipitation regime of a geographic region explains heterogeneity in incidence rates given climatic anomalies, we found that increasing summer temperatures were associated with especially pronounced relative increase in incidences among cooler counties as compared to already hot counties. At the same time, declines in winter precipitation were associated with strongly suppressed incidence in already very dry counties, but had a smaller effect on overall incidence in wetter counties. This finding, along with our estimates of drought-attributable cases across counties, suggests the presence of “ limiting factors” in the lifecycle of *Coccidioides* that vary by region. In arid regions, limited precipitation may restrict growth. In contrast, in cooler and wetter regions, the limiting factors may be insufficient heat to lyse the mycelia into individual arthroconidia and/or desiccate the soil to facilitate dust emissions, or excessive moisture for growth. This may explain why incidence rates have increased most dramatically in wetter and cooler counties, like the central coast counties, compared to the arid southern San Joaquin Valley counties [5].

Anticipated changes in California’s climate may continue to change the spatiotemporal distribution of coccidioidomycosis in the decades to come. Average precipitation over winter months is projected to see a modest increase [66] while precipitation in autumn and spring is projected to decrease [10, 67], which may enhance conditions favorable for *Coccidioides* wet growth period and dry dispersion period. At the same time, anthropogenic climate change is expected, with medium-high confidence, to increase the duration, intensity, and frequency of temperature-driven drought in California [8, 68]. Accordingly, coccidioidomycosis incidence may continue to expand into historically wetter and cooler regions, such as coastal California counties. Future analysis should consider how these relationships resolved in this work will affect the spatiotemporal distribution of coccidioidomycosis under anticipated climate regimes in the decades to come. Increased drought is expected to dramatically change the agricultural landscape in California, as well as agricultural occupations and occupation-related mobility patterns. Future work should also consider how climate and land use interact to produce risk for *Coccidioides* transmission as well as how changes in future occupational exposures may affect incidence. We establish the individual and joint effects of temperature and precipitation, both key drivers of soil moisture, and thus factors affecting agricultural drought. Future work may apply similar approaches to understand the complex relationship between coccidioidomycosis and modelled soil moisture products [69]. Soil moisture estimates may offer a more direct measurement of the water available to soil-dwelling microbes, by accounting for irrigation and transport of snow melt. Future work may also resolve whether meteorological drought (lack of precipitation) or hydrological drought (reduced streamflow and groundwater levels) results in similar associations with coccidioidomycosis incidence [9].

The assignment of cases to census tracts of residence, which may lead to exposure misclassification, is a key limitation of the study, even as nearby census tracts, where a case may have been exposed (e.g., during work, recreation, or travel), may experience similar environmental conditions as in the census tract of residence. By using census tract-level environmental and outcome data, our study offers an improvement over prior work analyzing county-level data on minimizing statistical bias induced by aggregating heterogeneous environmental and outcome data across broad geographic regions, what is known as the modifiable areal unit problem (MAUP). However, our study is still subject to bias from aggregation of heterogeneously distributed spatial phenomenon, particularly in larger census tracts. What is more, when aggregating cases, patients were assigned to the month of their estimated date of onset, which was estimated either by the patient’s own report, or, absent this, as the date of specimen collection. Therefore, the lag between a change in a climatic factor and its associated change in disease incidence also includes the incubation period for coccidioidomycosis, which varies between 7 and 21 days, and, for some patients, a lag between symptom onset and healthcare seeking, which is reported to be a median of 22 days [70], and a lag between healthcare seeking and testing. Some included cases may therefore have been exposed long before the September through November period, while some cases truly exposed in September through November may be excluded. This exposure misclassification may bias the associations towards the null, even as we were able to detect strong associations.

Our focus on incident cases in the months of September through November, which enabled us to parse out the influence of specific timing of wet and dry periods, limited our ability to draw conclusions about how precipitation and temperature affect incidence at other times of the year. Still, September through November captures the peak transmission for coccidioidomycosis and incidence between September through November is highly correlated with the number of cases in a transmission year.

Moreover, our ensemble models examined incidence throughout the entire year, and the findings align qualitatively with the regression models that were limited to incident cases in the months of September through November as both analyses found that incidence is suppressed during a transmission year with low precipitation, and a dry period before a wet period may amplify the usual transmission-enhancing effect of the wet period. Also, in modelling associations between climate variability and disease incidence, we do not control for factors that may lie on the causal pathway between temperature and incidence, such as near-surface winds or vegetation, even as they may play an important role in spore dispersal. Finally, our results pertain to California, where *C. immitis* dominates, and may not be reliably extrapolated to other endemic areas, such as Arizona, where *C. posadasii* prevails. While previous studies have largely focused on explaining the role of precipitation and temperature on the epidemiology of coccidioidomycosis in Arizona as compared to California, it remains to be seen how the effect of precipitation on coccidioidomycosis in Arizona is mediated by antecedent conditions, such as drought, or across regions with differing precipitation distributions.

While epidemiological studies are a powerful tool for relating climatic conditions to disease incidence and suggesting underling biological fungal responses driving patterns of disease, laboratory studies examining growth rate and spore production under experimentally modified moisture and temperature conditions are needed to validate the biological response of *Coccidioides* to various stimuli. Laboratory conditions may also seek to establish which soil microbes act as antagonists or symbionts for *Coccidioides*, and compare the range of conditions under which their survival is possible [53]. Large-scale field studies measuring the probability of *Coccidioides* detection under various climatic conditions can support these findings, while capturing and monitoring the complex microbial, plant, and animal diversity of the soil environment. Combinations of such studies are needed to validate or refute the “ grow and blow” hypothesis, the small mammal reservoir hypothesis, or the “ soil sterilization” hypothesis.

Our results offer the most in-depth treatment to date of the role of climatic factors on the transmission of coccidioidomycosis in California. Our ensemble modeling approach yielded a highly predictive model (r^2^ > 0.9) that captured interaction between drought and non-drought conditions and nonlinear dynamics of temperature and precipitation through the inclusion of semi-parametric and machine learning algorithms, while simultaneously avoiding model overfitting through the use of leave-out-one-year cross validation and inclusion of simple, parametric models. The results provide evidence to motivate inclusion of drought monitoring and seasonal climate forecasts into coccidioidomycosis surveillance efforts and provide usable timetables for coccidioidomycosis planning and prevention activities. For instance, prevention measures for coccidioidomycosis, including respirators for workers disturbing soil, wetting of soil before digging, and keeping car windows closed when driving through dusty areas, is needed to protect public health following droughts [71]. Increased awareness of the general public as well as health care providers of the heightened risk of coccidioidomycosis during wetter years, especially those following drought, may aid in faster diagnosis and treatment [72]. Nevertheless, prevention of coccidioidomycosis can be challenging and almost unavoidable in some circumstances (e.g., insufficient water for soil wetting, inability of wildland firefighters to wear N95s near active fires), increasing urgency to marshal resources towards ongoing vaccine development.

## Conclusion

Drought may displace and amplify coccidioidomycosis incidence in subsequent years. Beyond the effect of droughts, wet winters, particularly following dry years, combined with hot summers, increase coccidioidomycosis incidence. These findings have implications for the future of coccidioidomycosis in California, where warming temperatures and increased drought may further shift the burden of coccidioidomycosis towards the wetter coastal and northern San Joaquin Valley counties.

## Data Availability

Because protected health information is contained in the data used in this project, the raw data, or processed and linked data, drawn from California's infectious diseases reporting systems cannot be shared.

## Supplemental Text

### Justification of model choice

Our analysis makes use of several modelling techniques, which are each suited to obtain different target parameters of interest. Here, we describe the justification for each technique in greater detail.

### Multilevel distributed lag nonlinear models

#### County-specific models

We estimated relationships using a quasi-Poisson likelihood approach, with monthly cases as the outcome variable and the log of each census tract’s population as an offset term, so that model coefficients reflected the log incidence rate ratio. The primary exposure variables were lagged total precipitation and mean temperature. These were modeled with natural cubic spline functions of smoothed three-month averages, with lags spanning 1 to 36 months prior to estimated date of onset. For each of the 36 lags, we controlled for other lags every three (precipitation) or six (temperature) months. This approach allowed us to generate distributed lag models in which we examined the lagged effect of precipitation and temperature across all 36 months, while accounting for the historical precipitation and temperature history over the time period of interest in a way that avoided over fitting. To determine the location of knots for the cubic spline, we systematically varied the location of internal knots placed at precipitation or temperatures corresponding to average percentiles across counties [38], selecting the model that minimized the sum of Q-AIC across all counties, where the Q-AIC is a modification of the Akaike information criterion (AIC) for quasi-likelihood models [40]. We also included a natural cubic spline for soil type (percent sand), as it may be correlated with precipitation or temperature and the outcome but not on the causal pathway between the exposure and outcome (e.g., as vegetation might be). A cubic spline on year was also included to account for reporting and other secular trends not due to climate. Model formulas expressing *Y_i,t_*, the number of cases in census tract *i* in month *t*, for rainfall (equation 1) and temperature (equation 2) were:

For *j* ∈ [1, 36]:

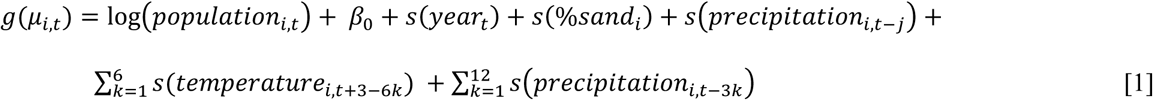

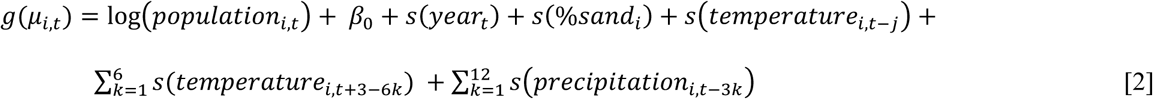

Where *g*() is a log function of the expectation *μ_i,t_* = *E*(*Y_i,t_*).

#### Pooled effects

Then, we used a fixed-effects meta-analysis to pool estimates of county-specific associations of precipitation and temperature with incidence [38]. We examined the overall shape of the associations between precipitation, temperature and incidence at each of the 36 monthly lags, and calculated the pooled IRR associated with an increase of precipitation or temperature from the 25^th^ percentile to the 75^th^ percentile (i.e., one interquartile range; IQR) over each lag.

Heterogeneity in the temperature-incidence and precipitation-incidence relationships across counties was additionally assessed using multivariate meta-regression models. This approach extends the fixed-effects meta-analysis to include meta-predictors, that summarize county-level information (e.g., mean county total precipitation, mean county temperature). The significance of the meta-predictors is determined using Wald tests [38]. For significant meta-predictors, we plotted separate exposure-response relationships across our set of meta-predictor values.

#### Effect modification

To examine potential effect modification of the wet winter period (which was determined by the fixed-effects meta-analysis to be the strongest predictor of fall coccidioidomycosis incidence) by antecedent conditions, we created a nonlinear interaction term between winter precipitation and antecedent conditions by multiplying the basis function for precipitation at a nine-month lag by a binary indicator for whether or not the census tract had a wetter or drier than average winter in the two years prior to exposure, three years prior to exposure, or both. The target parameter, the exponentiated coefficient on the modelled interaction term, is thus expressed as the ratio of the IRR for an IQR increase in total monthly winter precipitation following a dry year to the IRR for winter precipitation following a wet year. Deviations from mean were determined by calculating the percent difference of the monthly census tract value to the monthly county mean across the full time period examined. We used a single-stage distributed lag generalized additive model that adjusted for county as a fixed effect, and used cluster robust standard errors to adjust for the repeated observations at the census tract-level. All other model terms were the same as in the two stage analysis. The model formulas to examine effect modification by a drier than average winter two years ago, and two and three years ago, respectively, were:

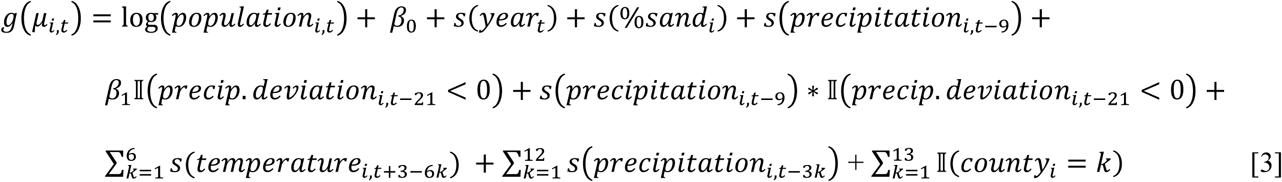

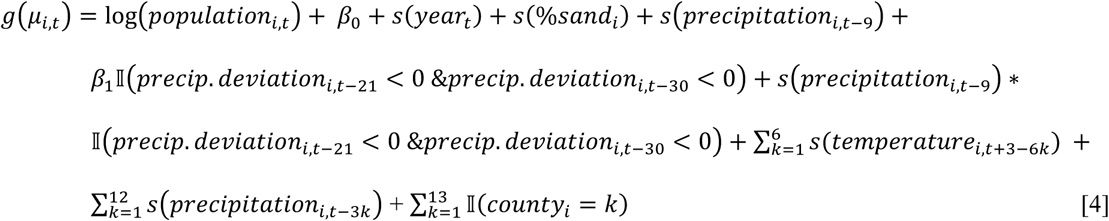

All statistical analyses were conducted in R, version 3.3.1 (R Foundation for Statistical Computing), using the splines and dlnm package for fitting distributed lag generalized additive models and the mvmeta package for performing fixed-effect meta-regression models [38, 41].

#### Ensemble modeling

We estimated cases attributable to—or averted because of—major droughts in California between 2000 and 2020 using a powerful, flexible ensemble modelling approach to predict incidence under counterfactual scenarios with regard to the presence or absence of drought [28]. For each county, we modelled the monthly incidence (for all months, January through February) at the census tract-level using five generalized linear models (GLMs) and one random forest algorithm. Table 2 summarizes the variables included in each of the six models. Model formulas expressing *Y_i,t_*, the number of cases in census tract *i* in month *t*, for the five GLMs were:

### Model 1

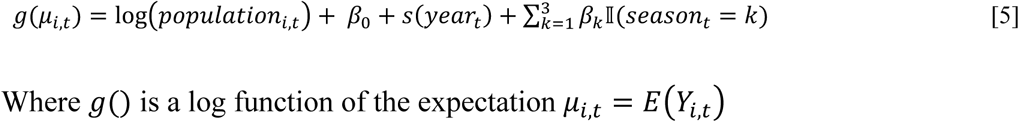

Where **g**() is a log function of the expectation μ*_i,t_* = *E*(*Y_i,t_*)

### Model 2

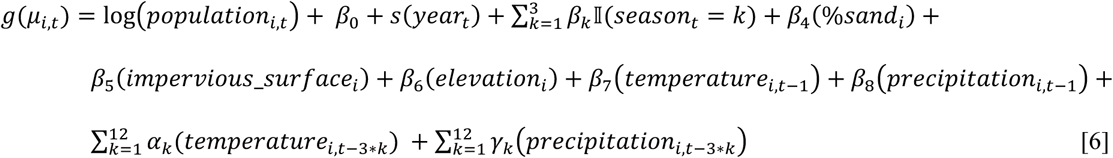

### Model 3

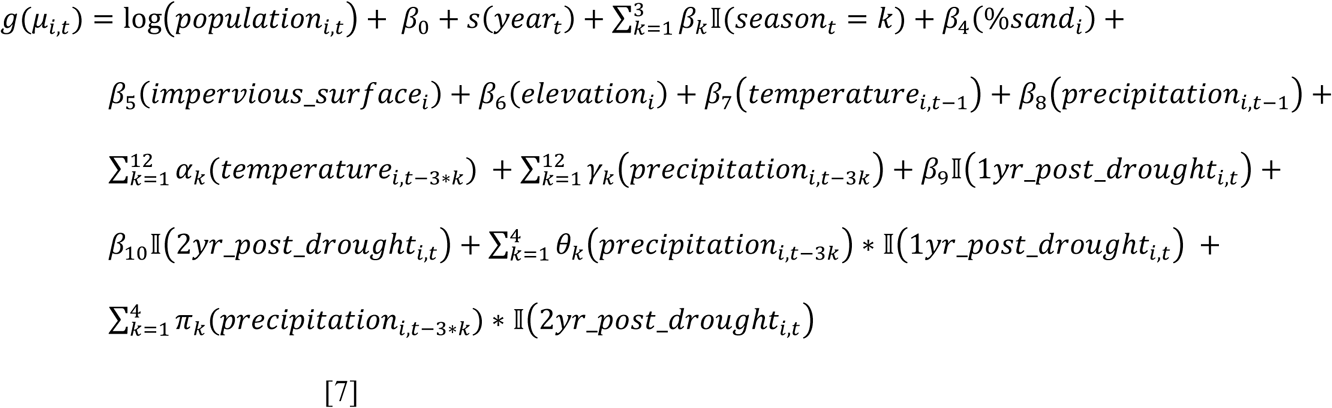

### Model 4

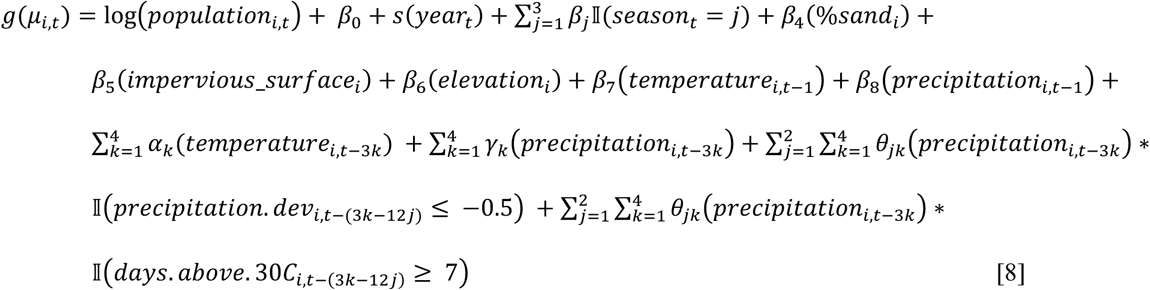

### Model 5

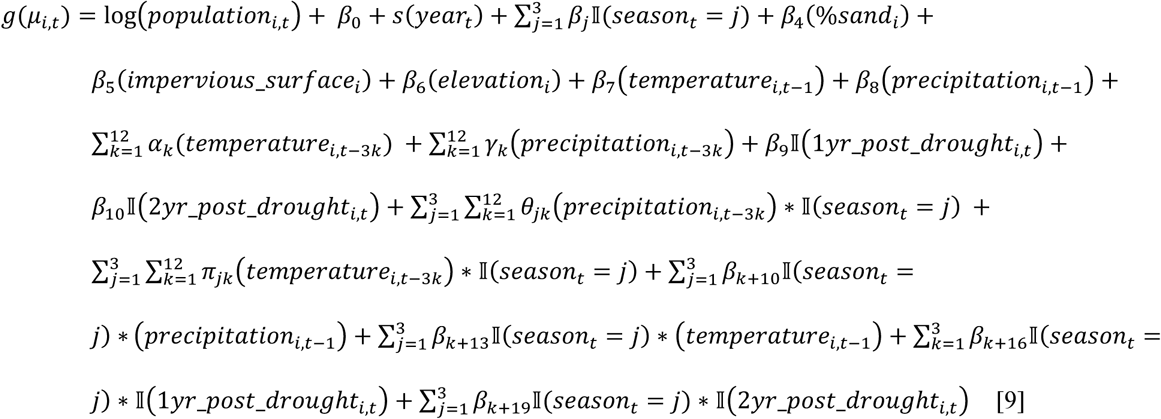

The sum of squared errors for each algorithm was calculated using leave-out-one-year cross-validation whereby the model was fit for all but one year of the time period, and then used to predict the out-of-sample cases in the left out year. We calculated a weight for each algorithm where the weights equaled the inverse of the cross-validated risk. Ensemble model predictions were then made for each census tract in each month as a weighted average of the individual model predictions.

We used the model to predict the total number of cases in a county over a certain time period under observed conditions, following the equation:

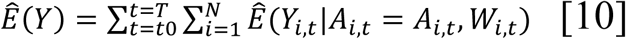

where:

**A_i,t_** represents the observed values of temperature and precipitation in census tract *i* at time *t*

**W_i,t_** represents the observed values for the other covariates in the model in census tract *i* at time *t*

*Y_i,t_* is the number of cases in census tract *i* at time *t* given observed values of temperature and precipitation and covariates

*N* is the total number of census tracts in the county

*t* ∈ (*t*0, *T*) represents the period of time of interest

The ensemble model was then used to predict the counterfactual number of cases, **Y**0, that would have been observed during the study period if, possibly contrary to what was observed, monthly average temperature higher than the historical average and total monthly precipitation below the historical average were deterministically set to their monthly county-level means.

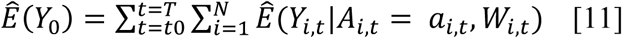

where:

*a_i,,t_* represents the counterfactual values for temperature and precipitation in census tract *i* at time *t*, obtained by setting any value for temperature above the mean equal to the mean, and any value for precipitation below the mean equal to the mean.

**W_i,t_** represents the observed values for the other covariates in the model in census tract *i* at time *t*

The incident cases attributable to—or averted by—the drought was estimated as the difference between predicted cases under the observed conditions and those predicted under the counterfactual “average climate” scenario. Because antecedent conditions as far back as three years may carry influence, we examined the attributable incidence during the drought and in the two years following the end of the drought. We estimated the effects separately for the severe drought spanning May 2012 until October 2015, and for the less severe drought from March 2007 to November 2009. We calculated the target parameter,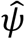, as follows, using equations [4] and [5]:

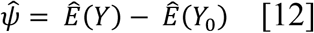

Because seasonality of coccidioidomycosis in California is such that incidence is lowest in March-April, with peaks occurring in the fall, we considered the change in incident cases “during drought” to include the period starting at the onset of the drought and extending until the end of the transmission season following the drought (e.g., March 31, 2010; March 31, 2016).

The two years post drought encompassed the full epidemiological seasons following the drought (e.g., April 1, 2010 – March 31, 2012; April 1, 2016 – March 31, 2018).

---

**Table S1.** Incidence rate ratios (IRRs) for fall coccidioidomycosis incidence associated with a one-unit increase in the IQR of temperature or precipitation

| **Month lag** | **IRR (95% CI) associated with a one unit increase in precipitation IQR** | **IQR of precipitation (mm)** | **IRR (95% CI) associated with a one unit increase in temperature IQR** | **IQR of temperature (°C)** |
| --- | --- | --- | --- | --- |
| 1 | 0.87 (0.80, 0.94) | [0.3, 11.2] | 1.29 (1.16, 1.44) | [18.1, 23.8] |
| 2 | 0.89 (0.85, 0.94) | [0, 2.5] | 1.55 (1.32, 1.82) | [20.6, 25.6] |
| 3 | 0.95 (0.92, 0.98) | [0, 2] | 2.02 (1.84, 2.22) | [20.3, 25.8] |
| 4 | 0.87 (0.81, 0.94) | [0.2, 8.3] | 0.58 (0.5, 0.66) | [18.3, 24.5] |
| 5 | 1.38 (1.29, 1.48) | [2.3, 20.7] | 0.49 (0.44, 0.54) | [15.9, 21.3] |
| 6 | 1.45 (1.36, 1.55) | [9.5, 41.1] | 0.5 (0.46, 0.55) | [13.6, 17.8] |
| 7 | 1.36 (1.28, 1.45) | [19.1, 57.4] | 0.58 (0.54, 0.63) | [11.7, 15.1] |
| 8 | 1.39 (1.29, 1.49) | [26.9, 71.3] | 0.57 (0.51, 0.64) | [10.1, 13.1] |
| 9 | 1.45 (1.36, 1.55) | [27.1, 73.2] | 0.74 (0.69, 0.79) | [9.5, 12] |
| 10 | 1.29 (1.20, 1.38) | [21.2, 64.4] | 0.83 (0.76, 0.91) | [9.6, 13.4] |
| 11 | 0.90 (0.84, 0.96) | [11.8, 44.4] | 0.98 (0.89, 1.07) | [11.6, 17.2] |
| 12 | 0.94 (0.87, 1.01) | [3.8, 24.7] | 1.11 (1.02, 1.2) | [14.4, 20.6] |
| 13 | 1.01 (0.94, 1.09) | [0.3, 11.2] | 1.23 (1.11, 1.35) | [18.1, 23.8] |
| 14 | 1.05 (1.01, 1.09) | [0, 2.5] | 1.29 (1.11, 1.5) | [20.6, 25.6] |
| 15 | 1.04 (1.00, 1.08) | [0, 2] | 1.16 (0.95, 1.41) | [20.3, 25.8] |
| 16 | 1.24 (1.15, 1.33) | [0.2, 8.3] | 0.79 (0.67, 0.94) | [18.3, 24.5] |
| 17 | 0.99 (0.93, 1.07) | [2.3, 20.7] | 0.77 (0.68, 0.88) | [15.9, 21.3] |
| 18 | 1.00 (0.93, 1.07) | [9.5, 41.1] | 0.8 (0.73, 0.88) | [13.6, 17.8] |
| 19 | 1.03 (0.96, 1.10) | [19.1, 57.4] | 0.87 (0.79, 0.95) | [11.7, 15.1] |
| 20 | 0.97 (0.92, 1.03) | [26.9, 71.3] | 0.88 (0.78, 1) | [10.1, 13.1] |
| 21 | 0.93 (0.88, 0.98) | [27.1, 73.2] | 1.35 (1.26, 1.44) | [9.5, 12] |
| 22 | 0.91 (0.86, 0.97) | [21.2, 64.4] | 1.31 (1.19, 1.43) | [9.6, 13.4] |
| 23 | 1.02 (0.95, 1.09) | [11.8, 44.4] | 1.22 (1.11, 1.34) | [11.6, 17.2] |
| 24 | 1.08 (1.01, 1.16) | [3.8, 24.7] | 1.2 (1.11, 1.31) | [14.4, 20.6] |
| 25 | 0.85 (0.79, 0.92) | [0.3, 11.2] | 1.28 (1.16, 1.41) | [18.1, 23.8] |
| 26 | 0.98 (0.94, 1.02) | [0, 2.5] | 1.24 (1.08, 1.43) | [20.6, 25.6] |
| 27 | 1.06 (1.01, 1.12) | [0, 2] | 1.24 (1, 1.54) | [20.3, 25.8] |
| 28 | 1.10 (1.02, 1.19) | [0.2, 8.3] | 1.05 (0.88, 1.25) | [18.3, 24.5] |
| 29 | 0.74 (0.69, 0.79) | [2.3, 20.7] | 0.99 (0.88, 1.12) | [15.9, 21.3] |
| 30 | 0.62 (0.58, 0.67) | [9.5, 41.1] | 1.03 (0.94, 1.12) | [13.6, 17.8] |
| 31 | 0.78 (0.73, 0.84) | [19.1, 57.4] | 0.98 (0.9, 1.06) | [11.7, 15.1] |
| 32 | 0.87 (0.82, 0.93) | [26.9, 71.3] | 1.03 (0.92, 1.15) | [10.1, 13.1] |
| 33 | 0.81 (0.77, 0.86) | [27.1, 73.2] | 0.99 (0.92, 1.06) | [9.5, 12] |
| 34 | 0.80 (0.76, 0.85) | [21.2, 64.4] | 1.13 (1.04, 1.22) | [9.6, 13.4] |
| 35 | 0.90 (0.84, 0.96) | [11.8, 44.4] | 1.07 (0.98, 1.16) | [11.6, 17.2] |
| 36 | 0.90 (0.84, 0.96) | [3.8, 24.7] | 1.1 (1, 1.2) | [14.4, 20.6] |

**Figure S1.**
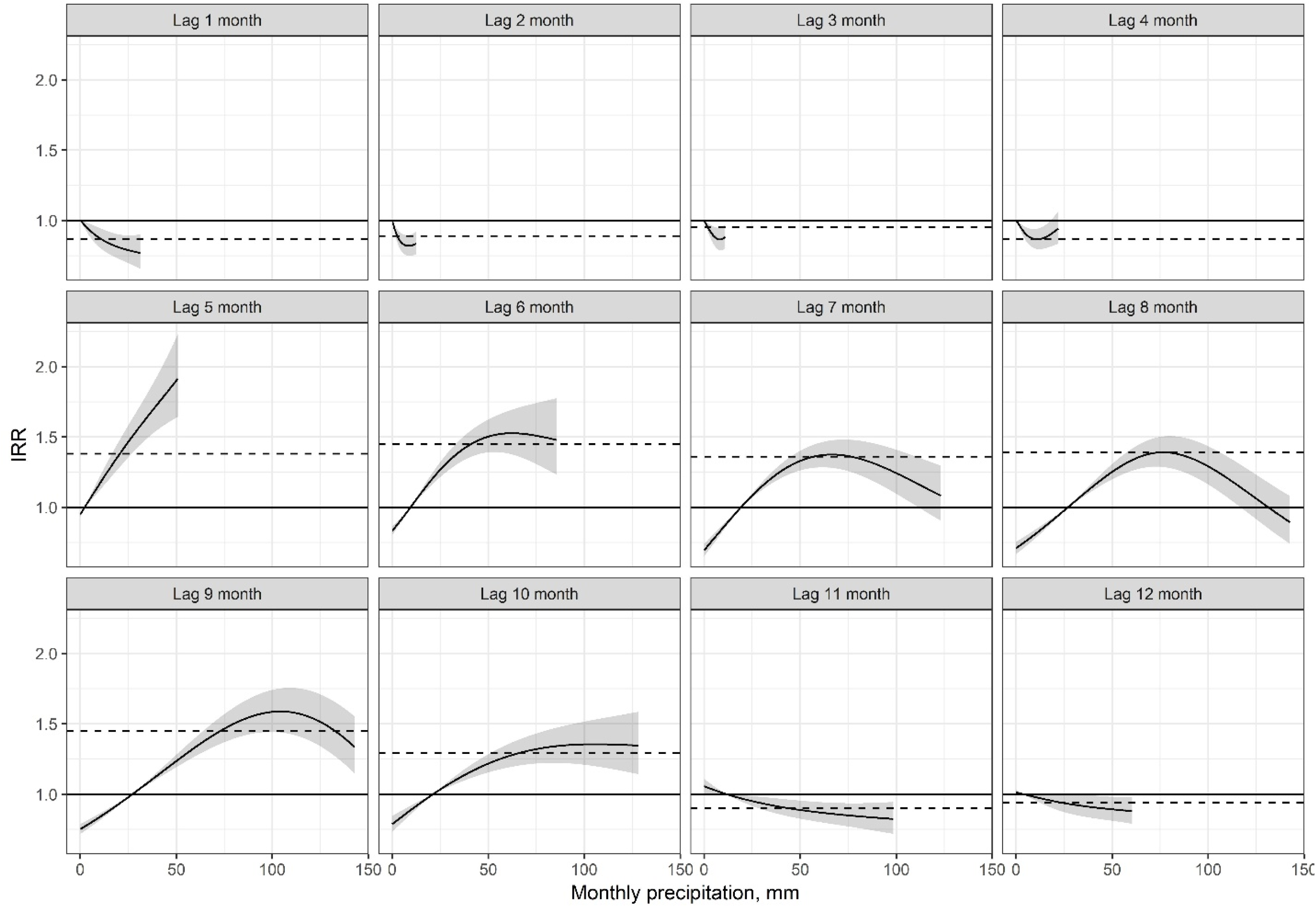
Each line represents a specific county, colored by their median annual precipitation. The lines depict where the *n*^th^ percentile of monthly total precipitation at a given lag falls along the pooled exposure-response relationship reflecting the relationship between lagged precipitation and coccidioidomycosis incidence. IRRs are scaled such that the reference is the 25^th^ percentile of precipitation at a given lag for the full study region.

**Figure S2.**
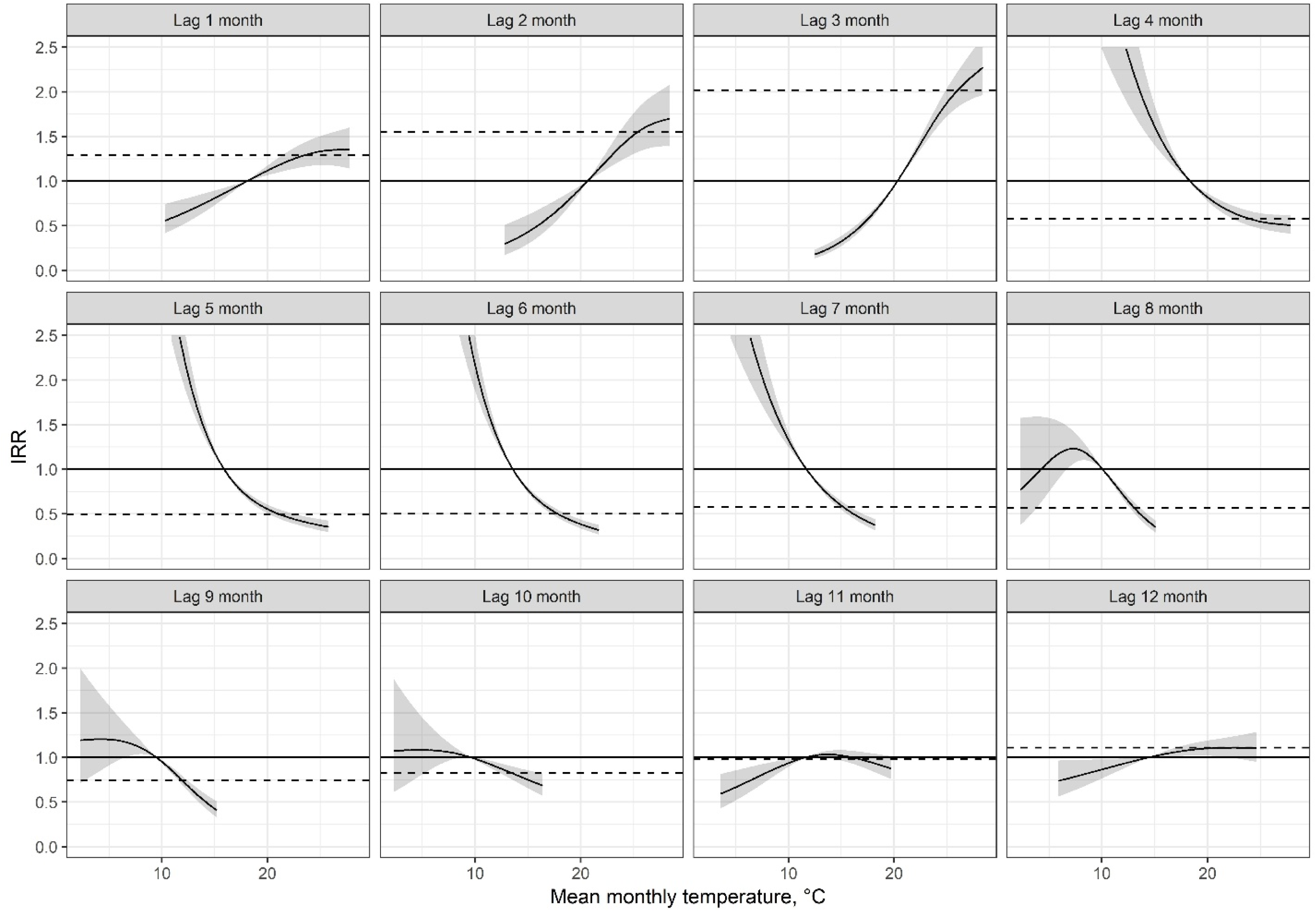
Each line represents a specific county, colored by their mean monthly temperatures. The lines depict where the *n*^th^ percentile of monthly temperature at a given lag falls along the pooled exposure-response relationship reflecting the relationship between lagged temperature and coccidioidomycosis incidence. IRRs are scaled such that the reference is the 25^th^ percentile of temperature at a given lag for the full study region.

**Figure S3.**
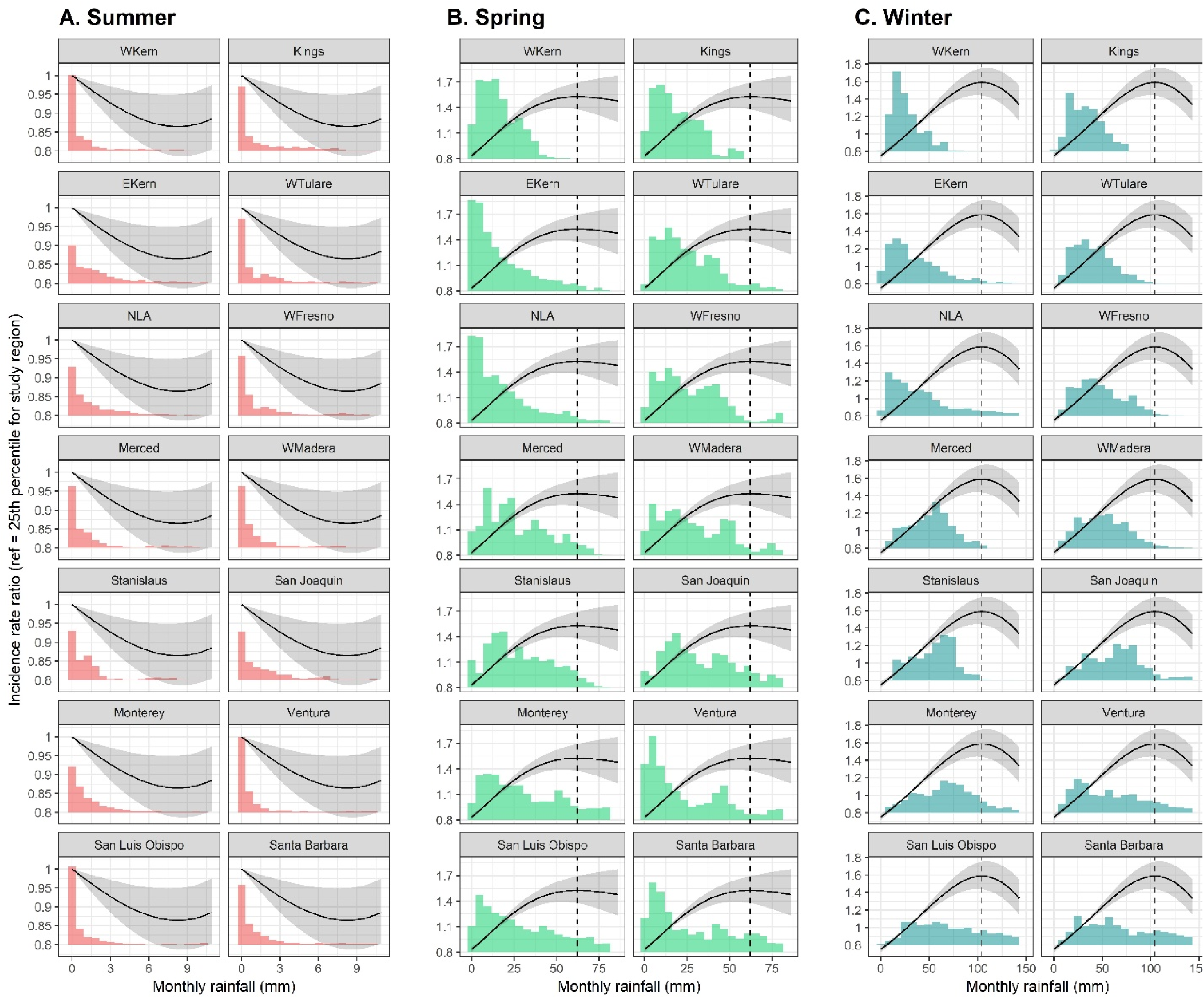
Pooled nonlinear relationship between change in temperature and precipitation at a 3, 6, and 9-month lags, corresponding to summer, spring, and winter, respectively. The IRR is centered at the 25^th^ percentile for the study region, such that the black curve indicates the incidence rate for a given temperature or precipitation value compared to the incidence rate at the 25^th^ percentile for the study region in that time period. Shaded gray regions reflect the 95% confidence interval. Precipitation corresponding to the maximum is indicated by the vertical dotted line. Histograms reflect the density of precipitation experienced by each county/sub-county in the study region.

**Figure S4.**
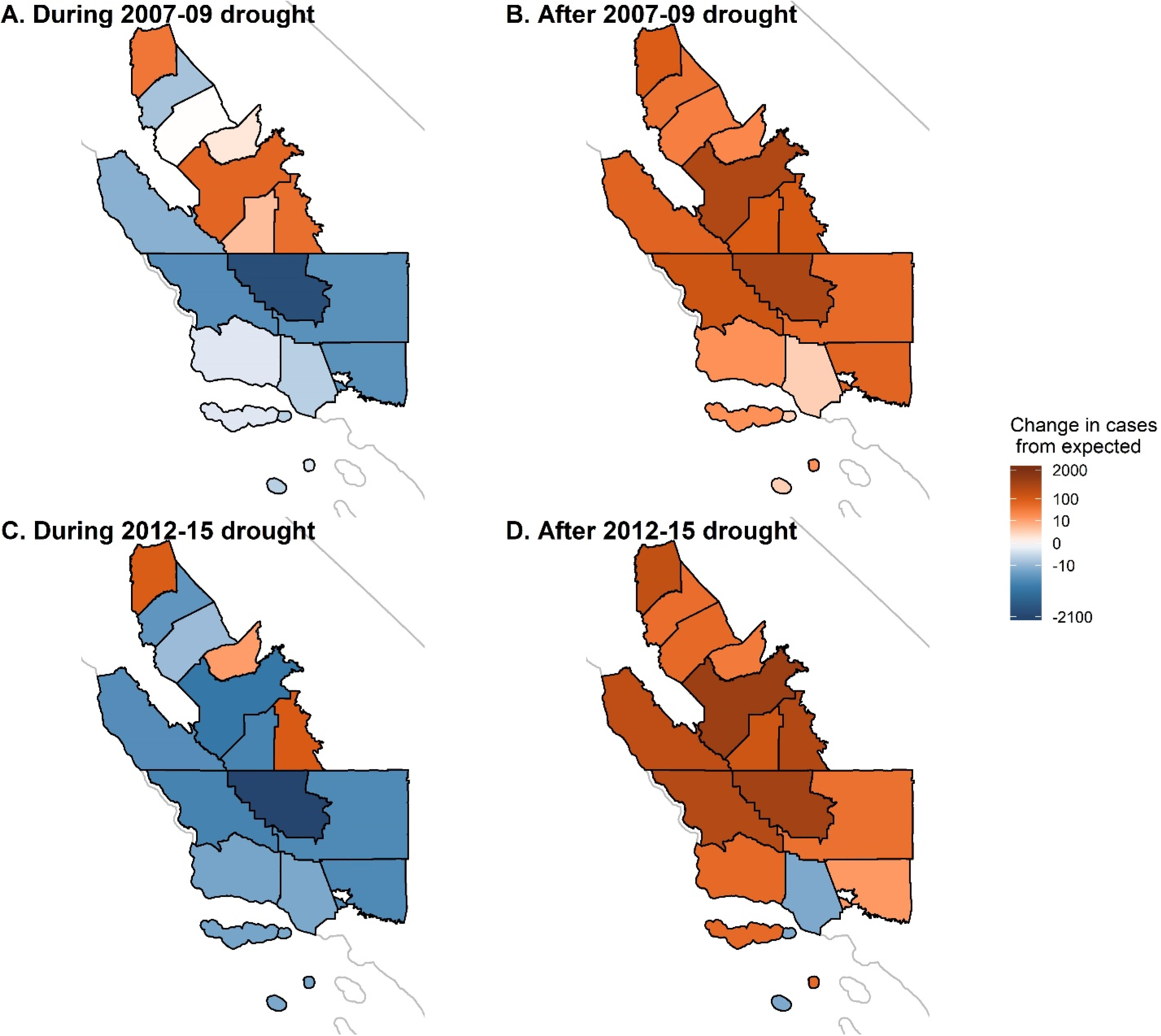
Estimated absolute excess (green) and averted (red) incident cases compared to the number expected in the absence of drought during (A and C) and in the two years following (B and D) the 2007-09 (A, B) and 2012-15 (C, D) droughts across the 14 counties in the study region. California state outline shown in light gray.

**Figure S5-18.**
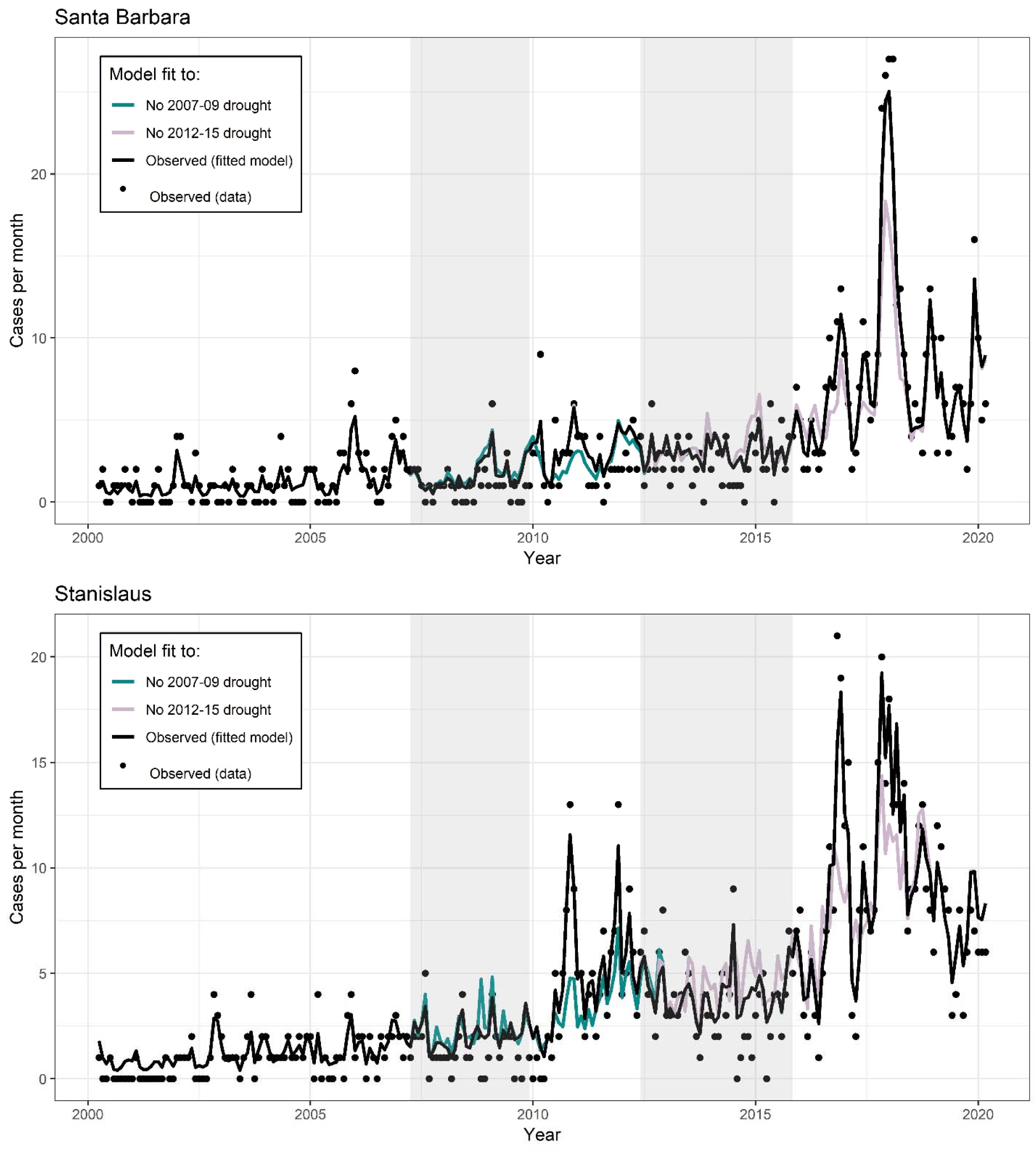
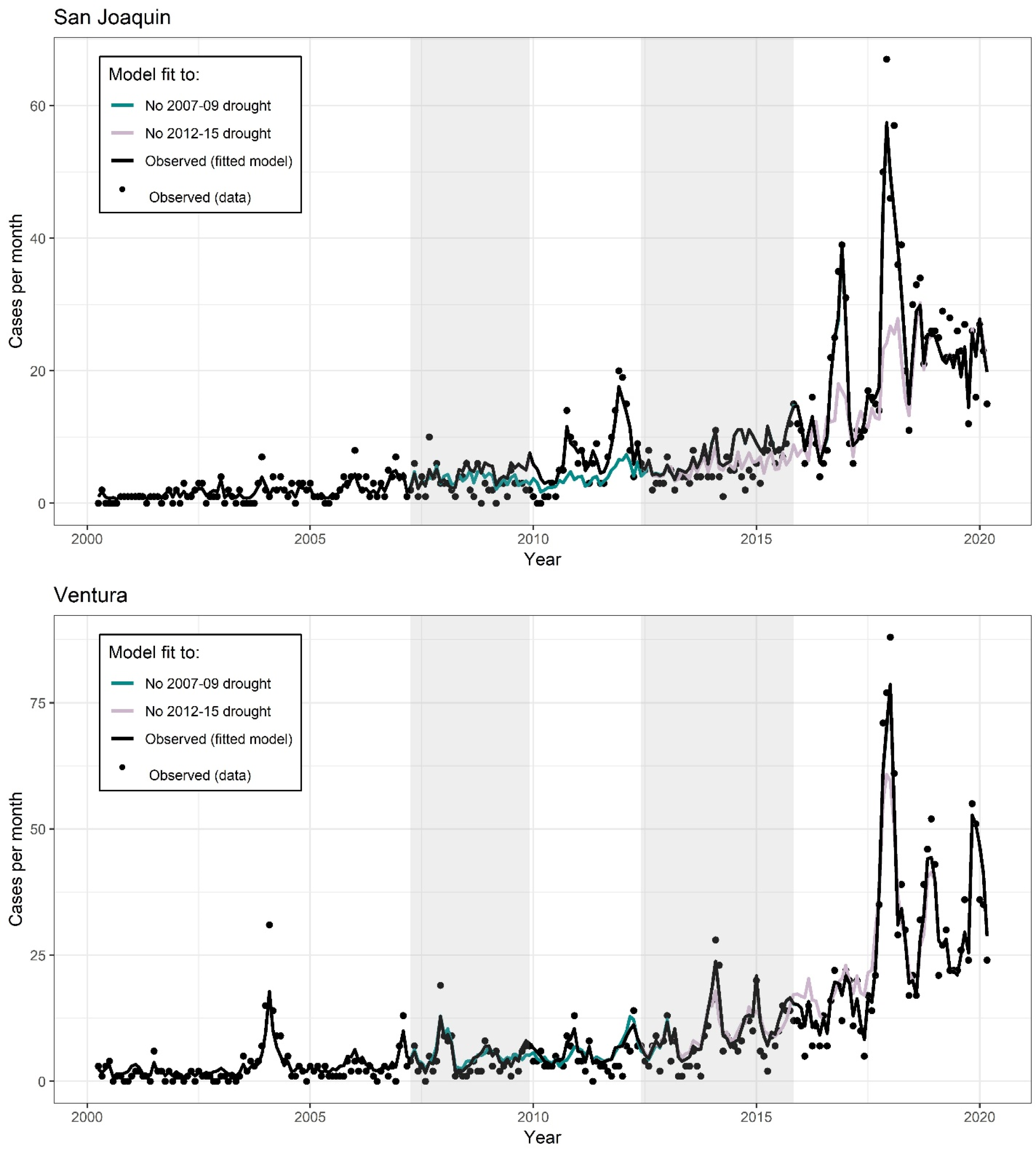
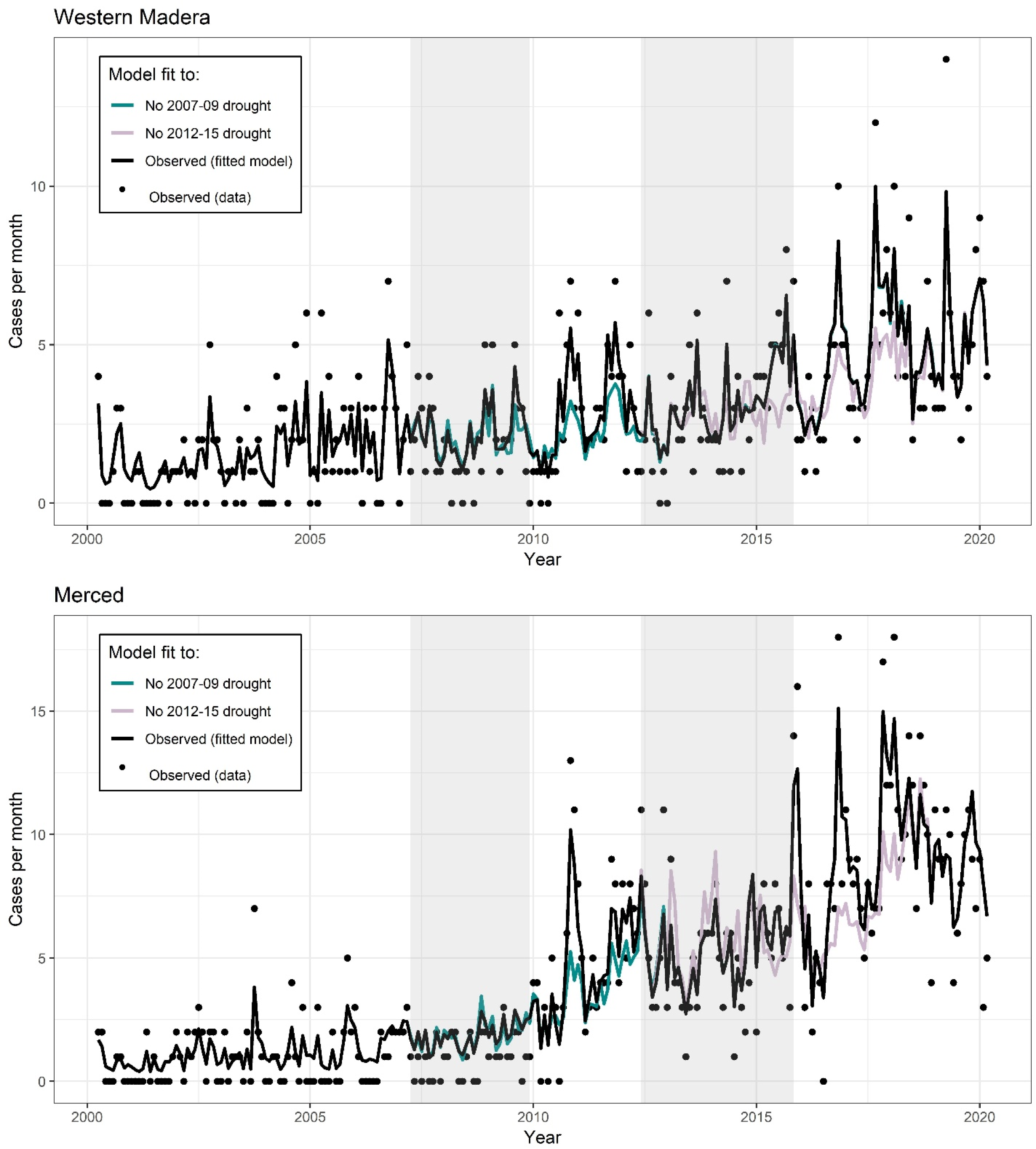
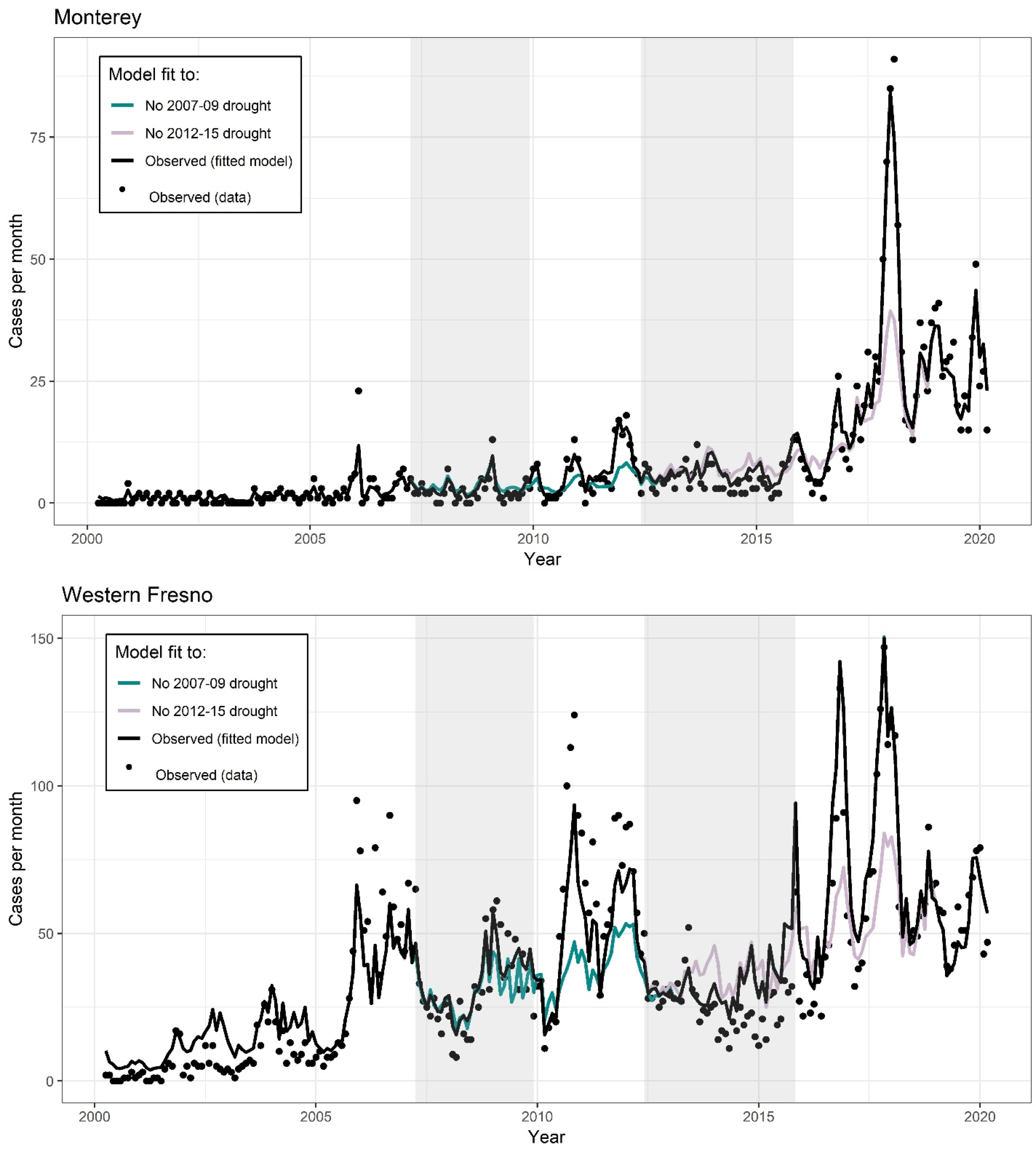
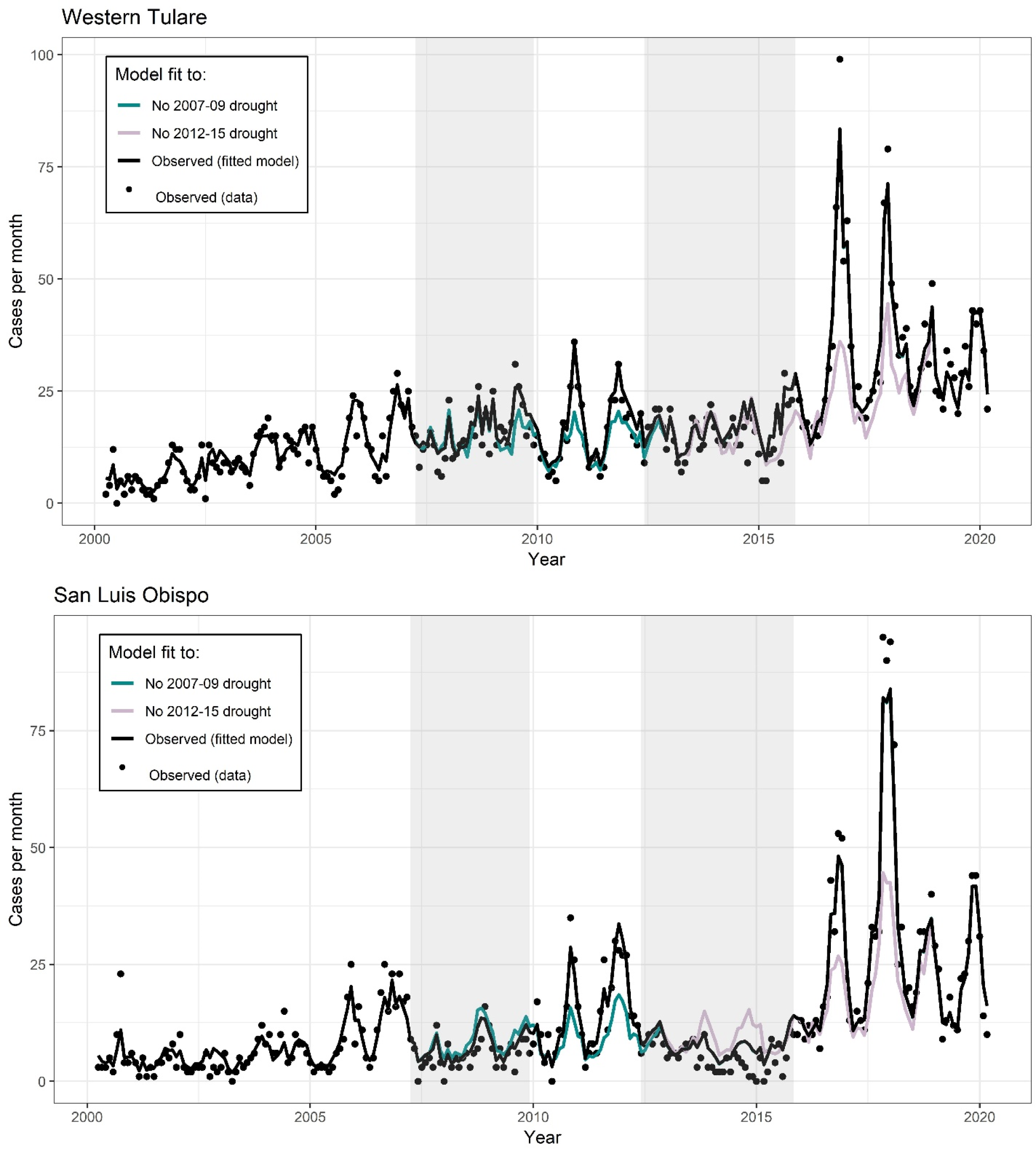
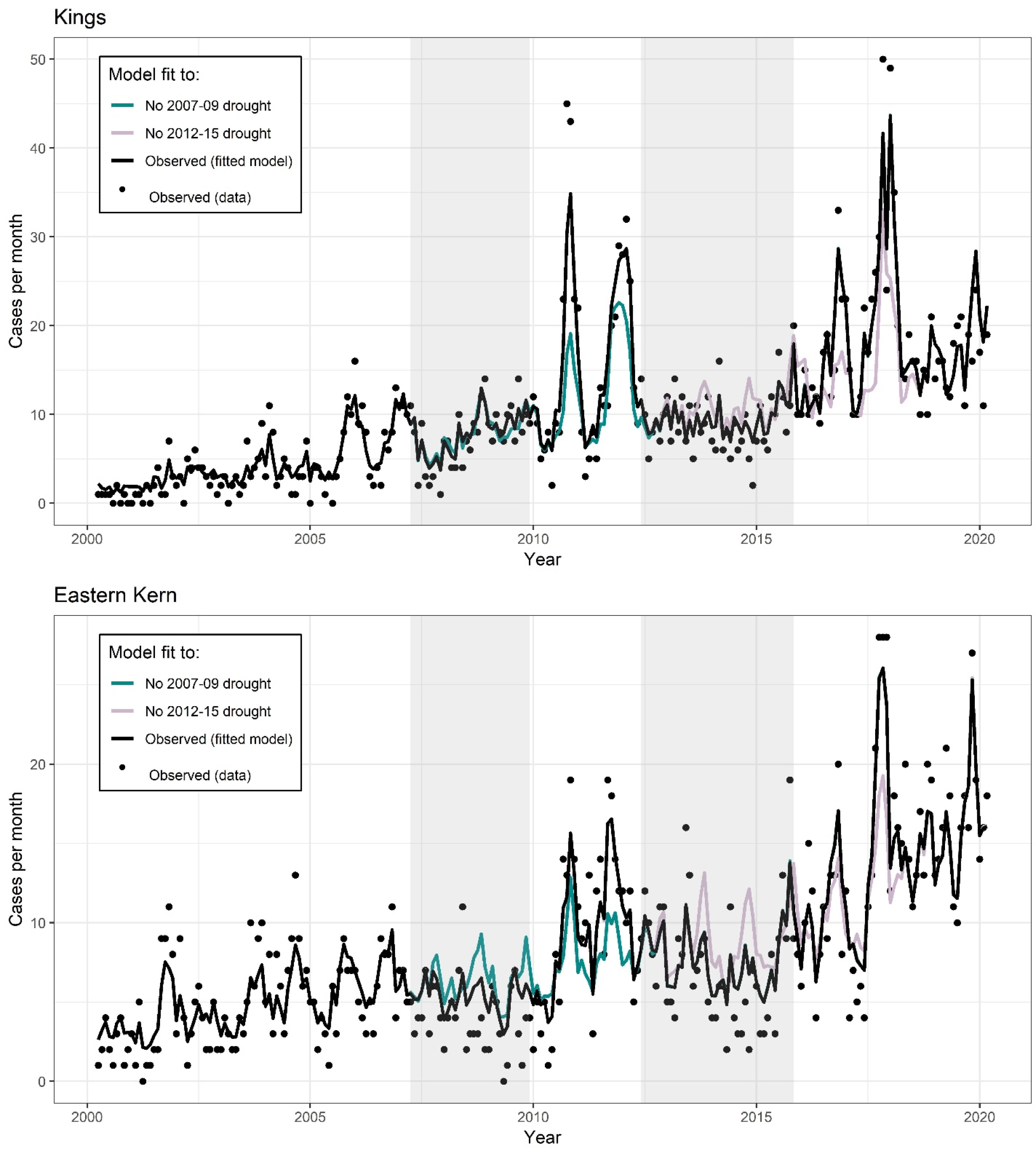
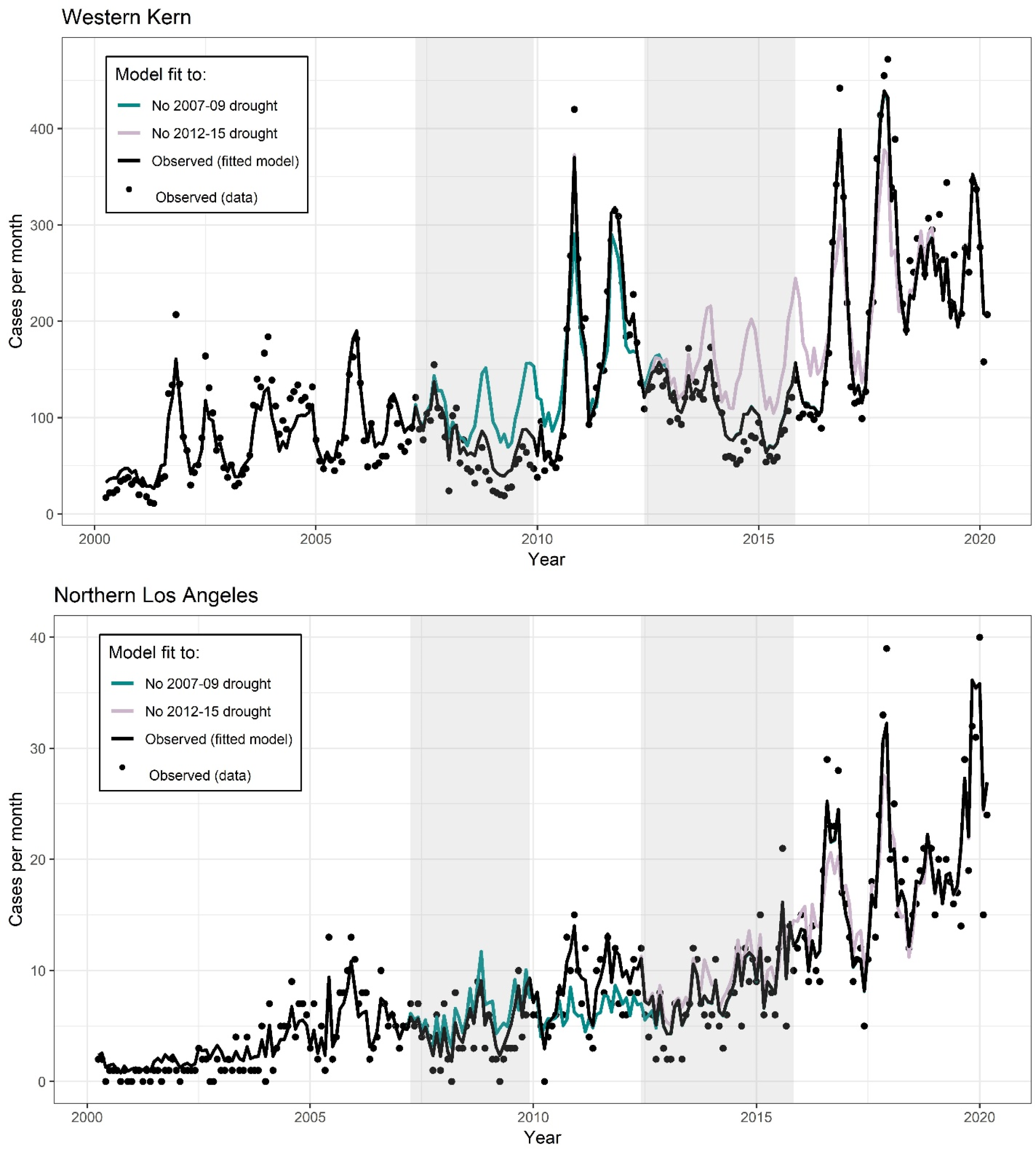
Observed incidence (black dots) by month for each county in the study region. Black line is the model fit under the observed environmental conditions. Color lines represent the expected incidence under the counterfactual intervention if the 2007-09 drought did not occur (cyan) or the 2012-15 drought did not occur (pink). Counterfactual scenarios were generated by setting temperatures observed to be higher than historical averages, and precipitation values observed to be below historical averages, deterministically to their average values.

## References

1. Galgiani JN, Ampel NM, Blair JE, Catanzaro A, Johnson RH, Stevens DA, et al. Coccidioidomycosis. Clin Infect Dis. 2005;41(9):1217–23.

2. Tabnak F LL, Sondermeyer G, Vugia D. . Epidemiologic Summary of Coccidioidomycosis in California, 2009-2012. Richmond, CA: California Department of Public Health; 2014.

3. Brown J, Benedict K, Park BJ, Thompson GR, 3rd. Coccidioidomycosis: epidemiology. Clinical epidemiology. 2013;5:185–97.

4. Stevens DA. Coccidioidomycosis. New England Journal of Medicine. 1995;332(16):1077–82.

5. Cooksey GLS, Nguyen A, Vugia D, Jain S. Regional Analysis of Coccidioidomycosis Incidence—California, 2000–2018. Morbidity and Mortality Weekly Report. 2020;69(48):1817.

6. Stewart ER, Thompson GR. Update on the Epidemiology of Coccidioidomycosis. Current Fungal Infection Reports. 2016;10(4):141–6.

7. National Center for Atmospheric Research Staff (Eds). The Climate Data Guide: PRISM High-Resolution Spatial Climate Data for the United States: Max/min temp, dewpoint, precipitation 2017 [Available from: https://climatedataguide.ucar.edu/climate-data/prism-high-resolution-spatial-climate-data-united-states-maxmin-temp-dewpoint.

8. Diffenbaugh NS, Swain DL, Touma D. Anthropogenic warming has increased drought risk in California. Proceedings of the National Academy of Sciences. 2015;112(13):3931–6.

9. United States Geologic Service. California Drought: What is drought? : USGS; 2021 [Available from: https://ca.water.usgs.gov/california-drought/what-is-drought.html.

10. Swain DL, Tsiang M, Haugen M, Singh D, Charland A, Rajaratnam B, et al. The extraordinary California drought of 2013/2014: Character, context, and the role of climate change. Bull Am Meteorol Soc. 2014;95(9):S3–S7.

11. Griffin D, Anchukaitis KJ. How unusual is the 2012–2014 California drought? Geophysical Research Letters. 2014;41(24):9017–23.

12. Mao Y, Nijssen B, Lettenmaier DP. Is climate change implicated in the 2013–2014 California drought? A hydrologic perspective. Geophysical Research Letters. 2015;42(8):2805–13.

13. United States Department of Agriculture. Unites States Drought Monitor 2020 [Available from: https://droughtmonitor.unl.edu/.

14. Centers for Disease Control and Prevention. Valley Fever Maps Atlanta, GA: CDC; 2019 [Available from: https://www.cdc.gov/fungal/diseases/coccidioidomycosis/maps.html.

15. Gorris ME, Treseder KK, Zender CS, Randerson JT. Expansion of Coccidioidomycosis Endemic Regions in the United States in Response to Climate Change. GeoHealth. 2019;3(10):308–27.

16. Fisher FS, Bultman MW, Johnson SM, Pappagianis D, Zaborsky E. Coccidioides niches and habitat parameters in the southwestern United States: a matter of scale. Annals of the New York Academy of Sciences. 2007;1111:47–72.

17. Maddy KT. Ecological factors of the geographic distribution of Coccidioides immitis. Journal of the American Veterinary Medical Association. 1957;130(11):475–6.

18. Komatsu K, Vaz V, McRill C, Colman T. Increase in coccidioidomycosis-Arizona, 1998-2001. Jama. 2003;289(12):1500-.

19. Comrie AC. Climate factors influencing coccidioidomycosis seasonality and outbreaks. Environ Health Perspect. 2005;113(6):688–92.

20. Kolivras KN, Comrie AC. Modeling valley fever (coccidioidomycosis) incidence on the basis of climate conditions. Int J Biometeorol. 2003;47(2):87–101.

21. Weaver EA, Kolivras KN. Investigating the Relationship Between Climate and Valley Fever (Coccidioidomycosis). EcoHealth. 2018;15(4):840–52.

22. Gorris M, Cat L, Zender C, Treseder K, Randerson J. Coccidioidomycosis dynamics in relation to climate in the southwestern United States. GeoHealth. 2018;2(1):6–24.

23. Coopersmith EJ, Bell JE, Benedict K, Shriber J, McCotter O, Cosh MH. Relating coccidioidomycosis (valley fever) incidence to soil moisture conditions. GeoHealth. 2017;1:51–63.

24. Tamerius JD, Comrie AC. Coccidioidomycosis incidence in Arizona predicted by seasonal precipitation. PloS one. 2011;6(6):e21009.

25. Zender CS, Talamantes J. Climate controls on valley fever incidence in Kern County, California. Int J Biometeorol. 2006;50(3):174–82.

26. Talamantes J, Behseta S, Zender CS. Fluctuations in climate and incidence of coccidioidomycosis in Kern County, California: a review. Annals of the New York Academy of Sciences. 2007;1111:73–82.

27. Comrie AC, Glueck MF. Assessment of climate-coccidioidomycosis model: model sensitivity for assessing climatologic effects on the risk of acquiring coccidioidomycosis. Annals of the New York Academy of Sciences. 2007;1111:83–95.

28. Robins J. A new approach to causal inference in mortality studies with a sustained exposure period—application to control of the healthy worker survivor effect. Mathematical Modelling. 1986;7(9):1393–512.

29. Azar A, Smith K. Epidemiologic Summary of Coccidioidomycosis in California, 2017. Sacramento, California: California Department of Public Health; 2017.

30. California Department of Public Health. Coccidioidomycosis in California Provisional Monthly Report January - December 2018. Sacramento, CA: CDPH; 2018.

31. Centers for Disease Control and Prevention. Case definitions for infectious conditions under public health surveillance, coccidioidomycosis. 2008.

32. California Department of Public Health. CDPH IDB Guidance for Managing Select Communicable Diseases: Coccidioidomycosis Sacramento: CDPH; 2018 [Available from: https://www.cdph.ca.gov/Programs/CID/DCDC/CDPH%20Document%20Library/IDBGuidanceforCALHJs-Cocci.pdf.

33. ESRI. ArcGIS Server REST API: Release 10. In: Institute ESR, editor. Redlands, CA: Environmental Systems Research Institute; 2020.

34. Chaney NW, Wood EF, McBratney AB, Hempel JW, Nauman TW, Brungard CW, et al. POLARIS: A 30-meter probabilistic soil series map of the contiguous United States. Geoderma. 2016;274:54–67.

35. Multi Resolution Land Characteristics Consortium. National Land Cover Database. 2011.

36. United States Geologic Service. USGS National Elevation Database 2018 [Available from: https://catalog.data.gov/dataset/usgs-national-elevation-dataset-ned.

37. Guo C, Yang J, Guo Y, Ou Q-Q, Shen S-Q, Ou C-Q, et al. Short-term effects of meteorological factors on pediatric hand, foot, and mouth disease in Guangdong, China: a multi-city time-series analysis. BMC infectious diseases. 2016;16(1):524.

38. Gasparrini A, Armstrong B, Kenward MG. Multivariate meta-analysis for non-linear and other multi-parameter associations. Statistics in medicine. 2012;31(29):3821–39.

39. Liu C, Yin P, Chen R, Meng X, Wang L, Niu Y, et al. Ambient carbon monoxide and cardiovascular mortality: a nationwide time-series analysis in 272 cities in China. The Lancet Planetary Health. 2018;2(1):e12–e8.

40. Peng RD, Dominici F, Louis TA. Model choice in time series studies of air pollution and mortality. Journal of the Royal Statistical Society: Series A (Statistics in Society). 2006;169(2):179–203.

41. R Core Team. R: A language and environment for statistical computing. Vienna, Austria: R Foundation for Statistical Computing; 2015.

42. Polley E, Van Der Laan M. Super Learner in prediction. UC Berkeley Division of Biostatistics Working Paper Series. Working Paper 266, May 2010, http://biostats.bepress.com/ucbbiostat/paper266; 2010.

43. Wright MN, Ziegler A. ranger: A Fast Implementation of Random Forests for High Dimensional Data in C++ and R. Journal of Statistical Software. 2017;77.i01.

44. Baumgardner DJ, Paretsky DP, Baeseman ZJ, Schreiber A. Effects of season and weather on blastomycosis in dogs: Northern Wisconsin, USA. Medical Mycology. 2011;49(1):49–55.

45. Greene DR, Koenig G, Fisher MC, Taylor JW. Soil isolation and molecular identification of Coccidioides immitis. Mycologia. 2000;92(3):406–10.

46. Barker BM, Tabor JA, Shubitz LF, Perrill R, Orbach MJ. Detection and phylogenetic analysis of Coccidioides posadasii in Arizona soil samples. Fungal Ecology. 2012;5(2):163–76.

47. Friedman L, Smith CE, Pappagianis D, Berman R. Survival of Coccidioides immitis under controlled conditions of temperature and humidity. American Journal of Public Health and the Nations Health. 1956;46(10):1317–24.

48. Crits-Christoph A, Robinson CK, Barnum T, Fricke WF, Davila AF, Jedynak B, et al. Colonization patterns of soil microbial communities in the Atacama Desert. Microbiome. 2013;1(1):1–13.

49. Lynch R, King A, Farías ME, Sowell P, Vitry C, Schmidt S. The potential for microbial life in the highest-elevation (> 6000 masl) mineral soils of the Atacama region. Journal of Geophysical Research: Biogeosciences. 2012;117(G2).

50. Sharpton TJ, Stajich JE, Rounsley SD, Gardner MJ, Wortman JR, Jordar VS, et al. Comparative genomic analyses of the human fungal pathogens Coccidioides and their relatives. Genome research. 2009;19(10):1722–31.

51. Kerr JR. Bacterial inhibition of fungal growth and pathogenicity. Microbial Ecology in Health and Disease. 1999;11(3):129–42.

52. Kathiravan MK, Salake AB, Chothe AS, Dudhe PB, Watode RP, Mukta MS, et al. The biology and chemistry of antifungal agents: A review. Bioorganic & Medicinal Chemistry. 2012;20(19):5678–98.

53. Lauer A, Baal JD, Mendes SD, Casimiro KN, Passaglia AK, Valenzuela AH, et al. Valley Fever on the Rise-Searching for Microbial Antagonists to the Fungal Pathogen Coccidioides immitis. Microorganisms. 2019;7(2):31.

54. Köberl M, Müller H, Ramadan EM, Berg G. Desert Farming Benefits from Microbial Potential in Arid Soils and Promotes Diversity and Plant Health. PloS one. 2011;6(9):e24452.

55. Del Rocío Reyes-Montes M, Pérez-Huitrón MA, Ocaña-Monroy JL, Frías-De-León MG, Martínez-Herrera E, Arenas R, et al. The habitat of Coccidioides spp. and the role of animals as reservoirs and disseminators in nature. BMC infectious diseases. 2016;16(1):550-.

56. Taylor JW, Barker BM. The endozoan, small-mammal reservoir hypothesis and the life cycle of Coccidioides species. Medical Mycology. 2019;57(Supplement_1):S16–S20.

57. Emmons CW, Ashburn LL. The Isolation of Haplosporangium parvum n. sp. and Coccidioides immitis from Wild Rodents. Their Relationship to Coccidioidomycosis. Public Health Reports (1896-1970). 1942;57(46):1715–27.

58. Emmons CW. Coccidioidomycosis in Wild Rodents. A Method of Determining the Extent of Endemic Areas. Public Health Reports (1896-1970). 1943;58(1):1–5.

59. Kollath DR, Teixeira MM, Funke A, Miller KJ, Barker BM. Investigating the Role of Animal Burrows on the Ecology and Distribution of Coccidioides spp. in Arizona Soils. Mycopathologia. 2020;185(1):145–59.

60. Ernest SKM, Brown JH, Parmenter RR. Rodents, Plants, and Precipitation: Spatial and Temporal Dynamics of Consumers and Resources. Oikos. 2000;88(3):470–82.

61. Prugh LR, Deguines N, Grinath JB, Suding KN, Bean WT, Stafford R, et al. Ecological winners and losers of extreme drought in California. Nature Climate Change. 2018;8(9):819–24.

62. Lacy GH, Swatek FE. Soil ecology of Coccidioides immitis at Amerindian middens in California. Applied microbiology. 1974;27(2):379–88.

63. Stephens RB, Rowe RJ. The underappreciated role of rodent generalists in fungal spore dispersal networks. Ecology. 2020;101(4):e02972.

64. Buckee C, Noor A, Sattenspiel L. Thinking clearly about social aspects of infectious disease transmission. Nature. 2021;595(7866):205–13.

65. Schrieks T, Botzen WJW, Wens M, Haer T, Aerts JCJH. Integrating Behavioral Theories in Agent-Based Models for Agricultural Drought Risk Assessments. Frontiers in Water. 2021;3(104).

66. Neelin JD, Langenbrunner B, Meyerson JE, Hall A, Berg N. California winter precipitation change under global warming in the Coupled Model Intercomparison Project phase 5 ensemble. Journal of Climate. 2013;26(17):6238–56.

67. Diffenbaugh NS, Giorgi F. Climate change hotspots in the CMIP5 global climate model ensemble. Climatic Change. 2012;114(3):813–22.

68. Bedsworth L, Cayan D, Franco G, Fisher L, Ziaja S. California’s Fourth Climate Change Assessment. 2019.

69. Liang X, Lettenmaier DP, Wood EF, Burges SJ. A simple hydrologically based model of land surface water and energy fluxes for general circulation models. Journal of Geophysical Research: Atmospheres. 1994;99(D7):14415–28.

70. Tsang CA, Anderson SM, Imholte SB, Erhart LM, Chen S, Park BJ, et al. Enhanced surveillance of coccidioidomycosis, Arizona, USA, 2007-2008. Emerg Infect Dis. 2010;16(11):1738–44.

71. California Department of Public Health. Valley Fever Prevention Tips Sacramento, CA: CDPH; 2021 [Available from: https://www.cdph.ca.gov/Programs/CID/DCDC/Pages/ValleyFeverPrevention.aspx.

72. Hurd-Kundeti G, Cooksey GLS, Jain S, Vugia DJ. Valley Fever (Coccidioidomycosis) Awareness—California, 2016–2017. Morbidity and Mortality Weekly Report. 2020;69(42):1512.

## References

1. Gasparrini A, Armstrong B, Kenward MG. Multivariate meta-analysis for non-linear and other multi-parameter associations. Statistics in medicine. 2012;31(29):3821-39.

2. Peng RD, Dominici F, Louis TA. Model choice in time series studies of air pollution and mortality. Journal of the Royal Statistical Society: Series A (Statistics in Society). 2006;169(2):179-203.

3. R Core Team. R: A language and environment for statistical computing. Vienna, Austria: R Foundation for Statistical Computing; 2015.

4. Robins J. A new approach to causal inference in mortality studies with a sustained exposure period—application to control of the healthy worker survivor effect. Mathematical Modelling. 1986;7(9):1393-512.

